# *α*-KIDS: A novel feature evaluation in the ultrahigh-dimensional right-censored setting, with application to Head and Neck Cancer

**DOI:** 10.1101/2024.08.13.24311946

**Authors:** Atika FArzana Urmi, Chenlu Ke, Dipankar Bandyopadhyay

## Abstract

Recent advances in sequencing technologies have allowed collection of massive genome-wide information that substantially enhances the diagnosis and prognosis of head and neck cancer. Identifying predictive markers for survival time is crucial for devising prognostic systems, and learning the underlying molecular driver of the cancer course. In this paper, we introduce *α*-KIDS, a model-free feature screening procedure with false discovery rate (FDR) control for ultrahigh dimensional right-censored data, which is robust against unknown censoring mechanisms. Specifically, our two-stage procedure initially selects a set of important features with a dual screening mechanism using nonparametric reproducing-kernel-based ANOVA statistics, followed by identifying a refined set (of features) under directional FDR control through a unified knockoff procedure. The finite sample properties of our method, and its novelty (in light of existing alternatives) are evaluated via simulation studies. Furthermore, we illustrate our methodology via application to a motivating right-censored head and neck (HN) cancer survival data derived from The Cancer Genome Atlas, with further validation on a similar HN cancer data from the Gene Expression Omnibus database. The methodology can be implemented via the R package DSFDRC, available in GitHub.

## 1. Introduction

Recent advancements in automated data collection techniques has led to an escalating prevalence of ultrahigh-dimensional data in biomedical sciences. Such data frequently exhibits an abundance of features that far surpasses the available number of observations. A salient example is evident in the massive data generated through high-throughput sequencing processes. With the ability to capture a wide spectrum of molecular, genetic, and phenotypic information on a large scale, researchers can now explore complex biological systems with unprecedented granularity and unveil novel insights into precision medicine, biomarker identification, and the exploration of pathways and networks. However, traditional statistical learning methods tend to falter when confronted with ultrahigh-dimensional data a predicament known as the ‘curse of dimensionality’. The present work was motivated by a study of the head and neck squamous cell carcinomas (HNSCC). Constituting around 4% of all cancer cases in the United States, head and neck cancer predominantly manifests as squamous cell carcinomas[1]. Despite surgery, radiation and chemotherapy, the 5-year survival rate stands at only 40-50% among all patients[30]. Extensive studies have found high biological and clinical heterogeneity in HNSCC patients, underscoring the need for a deeper molecular-level understanding of the disease.

Our goal in this paper is to identify which genes, among hundreds of thousands, contribute to survival prognosis of HNSCC, utilizing data from the HNSCC cohort available in the Cancer Genome Atlas (TCGA) network. The primary endpoint is the time to death (due to HNSCC). Due to loss to follow-up or no event occurrence until the study’s conclusion, more than half of cases were right-censored. As it is widely acknowledged that only a small subset of molecular features are truly relevant to specific clinical outcomes, feature selection has become one of the cornerstones for biomarker identification. While regularization methods such as LASSO [43], SCAD [13], adaptive Lasso [53], and Dantzig selector [6] have been the most popular feature selection tools, they can suffer from various issues including computational expediency, statistical inaccuracy and algorithmic instability [16] when applied to ultrahigh dimensional settings. A pragmatic solution is to perform feature screening before embarking on exact feature selection. A screening procedure applies a coarse filter to individual features to winnow out a significant portion of noise, thus circumventing a concurrent analysis of all features that gives rise to the aforementioned issues. This is frequently achieved through dependence learning between the outcome and individual features, which can be model-based [14, 23, 12], or model-free [52, 32, 35]. Given the challenges of verifying model assumptions in ultrahigh dimensional data, coupled with the risk of overlooking vital features due to model misspecification, opting for model-free screening approaches is a more prudent choice in practice.

In the regime of survival analysis, a plethora of model-free feature screening procedures have been developed for survival outcomes, subject to right censoring [40, 31, 51, 26, 7]. Since the true survival time is not fully observable, these approaches focus on the association between the estimated survival probabilities and individual features through, for examples, inverse-probability-of-censoring-weighted (IPCW) rank correlation [40], a generalized Kolmogorov statistic for covariate-stratified survival distribution [26], and distance correlation [7]. Nonetheless, there is no free lunch in estimating the survival function; assumptions on censoring are imposed, explicitly or implicitly, to ensure proper behavior of the survival estimators. The efficacy of the most widely used Kaplan-Meier estimation requires censoring to be independent of the event, as well as the predictive features for the survival time. Violations of independent censoring are not uncommon; subjects may tend to withdraw from the study due to either favorable or unfavorable prognosis, which can be further complicated by the effect of prognostic covariates. The ability of the IPCW method to adjust for dependent censoring hinges on the assumptions of exchangeability, and correct model specification used to estimate the weights. While a biased estimator undermines screening accuracy, it is extremely difficult to conjecture the censoring mechanism in practice to ensure unbiasedness, given ultrahigh dimensional covariates. Heavy censoring can also lead to less reliable estimators as the equivalent number of subjects at risk decreases at later times. The main impetus of this paper is the general lack of flexible and reliable screening tools for ultrahigh dimensional survival analysis, that are robust to heavy censoring and uncertain censoring mechanism as presented in the TCGA HNSCC dataset.

Moreover, in order to ensure the retention of important features with high confidence, feature screening procedures tend to opt for a conservative threshold to distinguish signal from noise. This approach, however, often leads to an excess of false discoveries. Consequently, it is essential to supplement the feature screening process with a more precise feature selection step to control the false discovery rate (FDR) in biomarker identification, thereby enhancing the accuracy of prognostic modeling.

Traditionally, feature selection and feature screening have been treated as separate domains in the literature. This division persisted until recent groundbreaking contributions in the form of the knockoff methodology [4, 5, 3, 33], which bridged this gap. The knockoff features are strategically designed to replicate the correlation structure inherent in the original variables, serving as negative controls to aid in the identification of truly significant features while controlling the FDR. This innovative approach can be extended to ultrahigh dimensional survival analysis, offering a solution to the longstanding challenge of FDR control in feature screening. In this paper, we propose a novel feature screening procedure with FDR control for ultra-high dimensional right-censored survival outcomes. The proposed method operates in two key stages. First, we find a preliminary set of potentially important features with a dual screening mechanism. Specifically, two filters are implemented to screen out irrelevant information through nonparametric dependence learning between the raw survival outcome and individual features, with no need for intermediate survival function estimation. The contribution of each feature is quantified by reproducing-kernel-based ANOVA statistics [28] in a model-free way. Then, we further identify a refined set of important features under directional FDR through a unified knockoff procedure based on the same utility measures adopted in the initial screening step. Our proposed method enjoys several distinctive advantages. First, it requires no pre-specification of the model structure and minimizes the assumption on the censoring mechanism. As a result, it exhibits more resilience to dependent and heavy censoring, than existing alternatives. Second, it effectively detects both linear and nonlinear features by capitalizing on the kernel-based utility measures. Third, it coherently controls FDR, and thus protects prognostic modeling from excessive noise. Finally, it boasts general applicability to other censored regression settings characterized by ultrahigh dimensional data with easy and fast implementation. All these advantages greatly facilitate the utilization of the proposed method in real applications. We substantiate both theoretically and numerically that the proposed feature evaluation procedure enjoys the sure screening property with rigorous control over FDR. Furthermore, through empirical analysis conducted on the TCGA HNSCC dataset and external validation, we showcase the efficacy and practical utility of the proposed method.

The rest of the paper is organized as follows. In Section 2, we develop our new framework of feature screening for ultrahigh dimensional survival analysis, and provide necessary theoretical justification. Simulation studies assessing finite sample properties of our proposal, and comparisons to existing alternatives are provided in Section 3. We illustrate our proposed methodology via application to the motivating TCGA HNSCC data in Section 4. Finally, Section 5 concludes, with a short discussion. All technical proofs, along with additional simulation results are deferred to the Web Supplement accompanying the paper.

## 2 Statistical Methods

In this section, we introduce a model-free dual screening framework for ultra-high dimensional right-censored data with FDR control. The proposed method is implemented via two main steps. First, it finds a crude set of potentially important features through a dual screening mechanism. Then, it further identifies a refined set of important features under directional FDR.

### 2.1 Assumptions and Dual Screening

Let *T* be the survival time and **X** = (*X*_1_, …, *X*_*p*_)^*T*^ be a p-dimensional set of covariates. We denote the censoring time by *C*. In reality, the observable survival response variables are (*Y, δ*), where *Y* = min*{T, C}* and *δ* = *I{T* ≤ *C}* with *I{·}* being the indicator function. Ideally, we would like to identify the smallest active set, denoted as 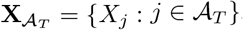, satisfying

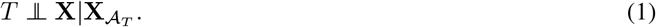

Since *T* is not fully observable, our focus shifts to identifying the smallest active set relevant to the observable outcome (*Y, δ*), denoted as **X**_*𝒜*_ = *{X*_*j*_ : *j* ∈ *𝒜}*, ensuring that

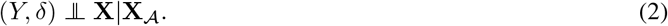

Nonetheless, the relation between the two active sets *A*_*T*_ and *A* can be established under a relatively simple condition, as shown in the following proposition.

#### Proposition 1

*Let* 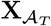 *and* **X**_*𝒜*_ *be the active sets that satisfy (1) and (2), respectively. Assuming that*

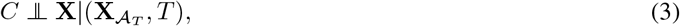

*we have* 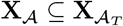.

The pair of conditions (1) and (3) is equivalent to (*T, C*) 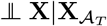, which mplies (*Y, δ*) **⊥** 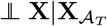 because (*Y, δ*) is a function of (*T, C*). It follows immediately that under condition (3), the important predictors for the observable outcome (*Y, δ*) are also important predictors for the true survival time *T*. Moreover, it is expected that in practice the equality 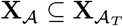 will normally hold since proper containment requires carefully balanced conditions. We note that assumption (3) is a mild condition since it allows censoring to vary with the true survival time and all the prognostic features. By contrast, the independent censoring condition,

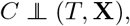

is more stringent and implies assumption (3). While many existing survival data screening methods assume independent censoring [40, 7, 10, 34], this assumption can be unrealistic in cases with complex censoring mechanisms and a large number of features. Another common assumption for ensuring identifiability [51, 26, 48],

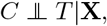

does not ensure the equivalence of 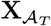 and **X** _*𝒜*_.

Throughout this paper, we assume that condition 3 holds, based on which a new framework of feature screening for right-censored data is developed. Specifically, we propose a new screening approach that directly applies to the raw survival outcome and thus avoids estimating the survival function. The following proposition lays the cornerstone for our method.

#### Proposition 2

*The pair of the following conditions* (*b*1) *and* (*b*2) *is equivalent to condition* (*a*):

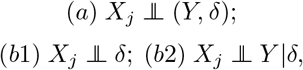

*for j* = 1, …, *p*.

According to Proposition 2, the pair of conditions (*b*1) and (*b*2) jointly implies the irrelevance of *X*_*j*_. Or, in other words, important features must be either marginally correlated with *δ* or conditionally correlated with *Y* given *δ*. Since *δ* is simply binary, the two conditions can be easily assessed by a wide range of independence measures. As a result, feature screening for right-censored data boils down to traditional univariate independence learning on complete data, bypassing the need to estimate the survival function. In this paper, we adopt a recently developed nonparametric independence measure, namely Expected Conditional Characteristic function Based Independence Criterion[28] (ECCFIC), as the filter in our screening procedure. We briefly review ECCFIC in the following subsection and illustrate its advantages over some existing celebrated independence measures.

### 2.2 Independence Measures

Let *U, V* ∈ ℝ be two random variables. Also, let *ϕ*_*U*_ denote the characteristic function of *U*, and *ϕ*_*U*|*V*_ denote the conditional characteristic function of *U* given *V*. Elicited by the fact that *U* **⊥** *V* if and only if *ϕ*_*U*|*V*_ = *ϕ*_*U*_, the ECCFIC for quantifying the association between *U* and *V* is defined by

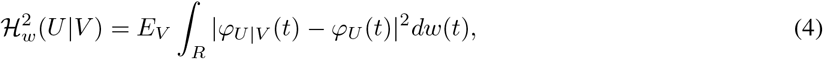

where, *w*(*·*) is a finite nonnegative Borel measure on R. An equivalent formula to (4) is given by

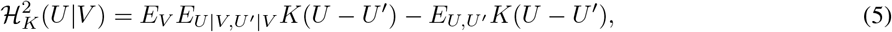

where (*U* ^*1*^, *V* ^*1*^) is an i.i.d. copy of (*U, V*), 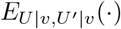 denotes conditional expectation *E*(*·*|*V* = *v, V* ^*1*^ = *v*), and *K* : ℝ → C is a translation-invariant positive definite kernel induced by *w*, such that *K*(*x*) = ∫_ℝ_ *e*^−*ixt*^*dw*(*t*) for *x* ∈ ℝ by Bochner Theorem [46]. Henceforth, we consider the alternative representation in (5), as it is easier to estimate for a given kernel. As a special case, we have

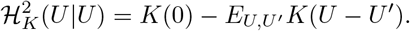

It can be shown that 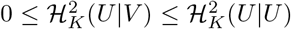. Moreover, if *K* is characteristic [19], then 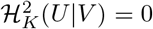 if and. only if *U* ⊥ *V*. As a result, we can define an *R*^2^-type statistic as

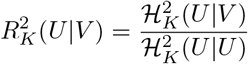

and 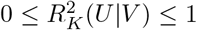. In particular, 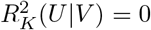 if and only if *U* **⊥** *V* and 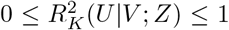 if and only if *U* is a measurable function of *V*. Intuitively, this kernel-based *R* statistic can be regarded as a nonlinear generalization of the classical *R*^2^ as it requires no linearity or distributional assumptions for the regression of *U* on *V*. It is worth noting that ECCFIC is closely related to a well-known family of measures, called Hilbert-Schmidt independence criterion[20] (HSIC), which includes distance covariances[42] as a special case. To assess the association between *U* and *V*, HSIC consider the discrepancy between the joint characteristic function *ϕ*_*U,V*_ and the product of the marginals *ϕ*_*U*_ *ϕ*_*V*_ [21], where the two random variables are treated symmetrically. Although HSIC equal zero also indicates independence and vice versa, it is not clear under what circumstances HSIC approaches its upper bound or how the random variables are related when the upper bound is attained. Compared to HSIC, ECCFIC better quantifies the contribution of a feature to the outcome, since it characterizes both independence and functional dependence in a supervised way, thereby making ECCFIC a more appealing alternative for model-free feature screening.

In the similar vein, the marginal effect associated with *V* (given another variable *Z* ∈ *ℝ* already contained in the model to explain *U*) can be measured by the expected conditional characteristic function based conditional independence criterion (ECCFCIC [28]):

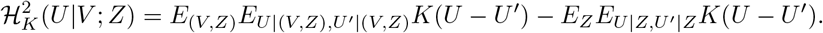

Then, a kernel-based partial *R*^2^ statistic can be defined by

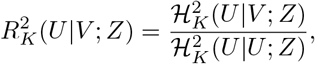

and 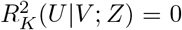 with 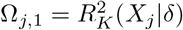 if and only if *U* **⊥** *V* |*Z*. Although here we restrict ourselves to translation-invariant kernel for ease of presentation, ECCFIC can be generalized using any positive definite characteristic kernel in the associated reproducing kernel Hilbert space[28]. Examples of characteristic kernel include but not limited to Gaussian, Laplacian, and inverse multiquadric.

### 2.3 The Screening Procedure

Returning to the context of feature screening for right-censored data, we propose to use the following two utility measures to evaluate each feature based on conditions (*b*1) and (*b*2), respectively:

1. 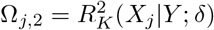
2. 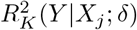

The first measure is the kernel-based *R*^2^ for the inverse regression of *X*_*j*_|*δ* and the second measure is the kernel-based partial *R*^2^ for the inverse regression of *X*_*j*_|*Y* while adjusting for *δ*. Here the inverse regression is to facilitate sample estimation as *δ* is a binary variable. We note again that any appropriate nonparametric dependence measures may be used to access conditions (*b*1) and (*b*2). For example, the marginal measure can be replaced by the Kolmogorov filter [35] or the MV-SIS filter [8], while the conditional measure can be replaced by 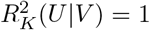 by exchanging *X*_*j*_ and *Y*. Given sample data *{*(**X**_*i*_, *Y*_*i*_, *δ*_*i*_) : *i* = 1, …, *n}*, we develop the estimators for the proposed utility measures. Let *𝒥*_*s*_ = *{i* : *δ*_*i*_ = *s}, n*_*s*_= |*𝒥*_*s*_|, and 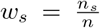 for *s* = 0, 1. Denote 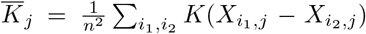 and 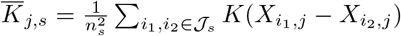 for *s* = 0, 1. The marginal utility can be estimated as

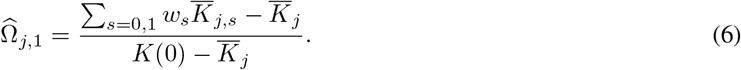

The conditional utility can be estimated as

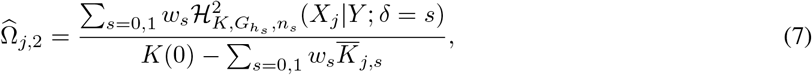

where, 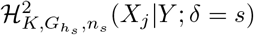 is the Nadaraya-Watson estimator of 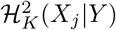 given *δ* = *s*, relying on a smoothing kernel *G* : ℝ → *ℝ* and an associated tuning bandwidth *h*_*s*_ = *h*_*s*_(*n*_*s*_) ∈ *ℝ*. Specifically,

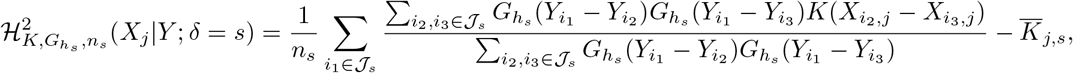

where,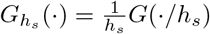.

According to Proposition 2, features making discernibly marginal or conditional contribution to the survival outcome should be retained. Therefore, we estimate the active index set by

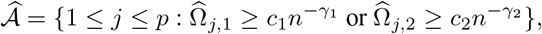

where, *c*_1_, *c*_2_, *γ*_1_ and *γ*_2_ are some threshold values relying on the strength of the true signal, which is to be defined in condition v below. Henceforth, we refer to the proposed screening procedure as *k*ernel-based *i*ndependence *d*ual *s*reening (abbreviated as KIDS). The proposed procedure embraces the sure screening property as well as the rank consistency property, which are established in Theorems 2.3 and 2.3 below.

Let *𝒜*_1_ = *{j* ∈ *𝒜* : *X*_*j*_ ⊥*/* ⊥ *δ}* and *𝒜*_2_ = *{j* ∈ *𝒜* : *X*_*j*_ ⊥*/* ⊥ *Y* |*δ}*. Then *𝒜* = *𝒜*_1_ ∪ *𝒜*_2_. The following regularity conditions are imposed to facilitate the technical proof, although, they may not be the weakest one.

libel=(C0),itemsep=0pt The characteristic kernel *K* is bounded.

liibel=(C0),iitemsep=0pt The smoothing kernel *G* : ℝ→ ℝ satisfies ∫_ℝ_ *y*^*i*^*G*(*y*)*dy* = *I{i* = 0*}* for *i* = 0 and 1, and *G*(*y*) = *O*((1 + |*y*|^4^)^−1^).

liiibel=(C0),iiitemsep=0pt *h*_*s*_ → 0 and 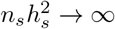 as *n*_*s*_ → ∞, for *s* = 0, 1.

livbel=(C0),ivtemsep=0pt The density of *Y* given *δ* = *s*, denoted as *f*_*Y,s*_(*y*), is bounded away from zero, for *s* = 0, 1. In addition, the first partial derivatives of *f*_*Y,s*_(*y*) is uniformly bounded by some constant that does not depend on *y*, for *s* = 0, 1.

lvbel=(C0),vtemsep=0pt There exist *c*_1_, *c*_2_ *>* 0 and *γ*_1_, *γ*_2_ ∈ [0, 1*/*2), such that 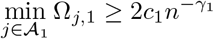 and 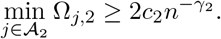.

lvibel=(C0),vitemsep=0pt There exist *c*_3_, *c*_4_ *>* 0 and *γ*_3_, *γ*_4_ ∈ [0, 1*/*2), such that 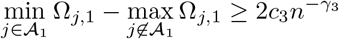 and 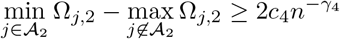.

Condition i is satisfied for many popular kernels [2]. Conditions ii-iv are commonly assumed for Nadaraya-Watson estimators. Condition v and vi are also standard in the literature of variable screening requiring that the true signal is detectable and is distinguishable from noise.

[Sure Screening] Under conditions i-v,

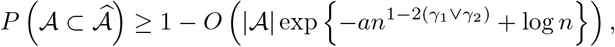

where, *a >* 0 is some constant.

[Rank Consistency] Under conditions i-iv and vi,

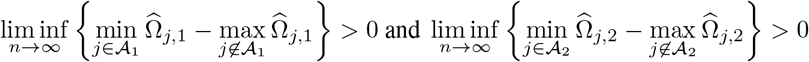

almost surely for 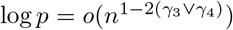.

Proofs of Theorems 2.3 and 2.3 appear in Web Supplement 6.2 and 6.3, respectively. Theorem 2.3 suggests that all the important features are selected asymptotically almost surely, and Theorem 2.3 further indicates that active features can be well separated from inactive ones. Both properties hold with NP-dimensionality 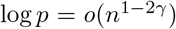 for some *γ* ∈ [0, 1*/*2).

There is no established way of determining the threshold values in a finite sample setting. As it is commonly assumed that the cardinality of the truly important set is small, one may specify a model size *d < n* and select 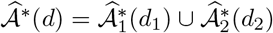, where

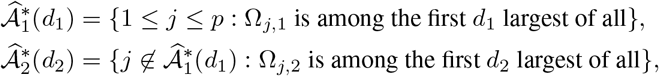

for *d*_1_ + *d*_2_ = *d*. Typical choices of *d* are [*n/* log(*n*)], 2[*n/* log(*n*)], 3[*n/* log(*n*)], and *n* − 1 [14, 32]. We can simply set *d*_1_ = *d*_2_ = [*d/*2], in which case the marginal and conditional utility measures are equally weighted in the selection of 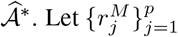 and 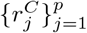 be the two rankings of variables by 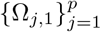 and 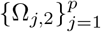, respectively. A joint ranking 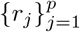 can be acquired by ascending 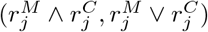. Then selecting the top *d* variables is identical to the trivial choice of 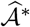 with *d*_1_ = *d*_2_. The sure screening property entails that the probability of selecting all the active predictors is close to one when *d* is sufficiently large. Inevitably, false discoveries can be inflated simultaneously with a generous choice of *d*. We address this issue in the next two subsections.

### 2.4 FDR Control via Knockoff

The most important assumption of ultrahigh dimensional problems is the sparsity principle, which assumes that the cardinality of 𝒜 is very small compared to *p*. In most cases, it is very hard, if not impossible, to recover *A* exactly without error. Ensuring all the active predictors are selected with high probability in the preceding screening procedure may introduce too much noise to the downstream analysis in the meanwhile. Therefore, a natural interest is to find a balancing trade-off between the sure screening property and the false discovery rate (FDR). In this subsection, we develop a dual selection procedure for right-censored data with FDR controlling using knockoff features.

We say 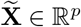 is a knockoff copy of **X** if

1. Swapping *X*_*j*_ with 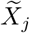 does not change the joint distribution of (**X**, 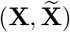);
2. 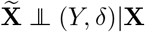.

The second condition is trivially achieved as long as 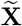 is constructed without using (*Y, δ*). However, if the distribution of **X** is unknown, how to obtain exact knockoff copies that satisfy the first condition remains elusive. Nonetheless, we may construct approximate second-order knockoff features, such that 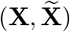 is pairwise exchangeable with respect to the first two moments. Suppose ***µ*** = *E*(**X**), Σ = *Cov*(**X**). Mean invariance can be easily achieved by forcing 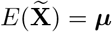. The second-order pairwise exchangeable condition is equivalent to

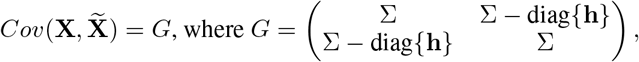

and **h** ∈ *ℝ*^*p*^ is a vector that makes *G* a positive semidefinite covariance matrix. Different approaches are available to select **h** [4]. For example, one may find **h** by solving

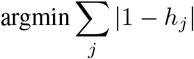

subject to *h*_*j*_ ≥ 0 and 2Σ − diag*{***h***}* being positive semidefinite. If we treat **X** as fixed [4] and normalize each feature such that the sample covariance 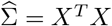 and 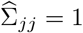 with *X* ∈ *ℝ*^*n×p*^ being the data matrix, then the knockoff data matrix 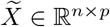 can be obtained by

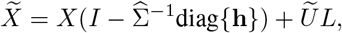

where, 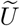 is an *n × p* orthonormal matrix that is orthogonal to the span of *X*, and 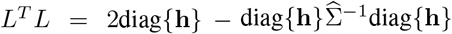 is a Cholesky decomposition. In a more general Model-**X** setting [5] where **X** has an unknown distribution, we can generate approximate knockoff features from conditional normal distribution as

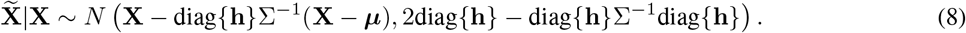

Note that, if **X** is Gaussian, the equivalence of the first two moments implies the equivalence of the joint distribution, such that (8) yields exact knockoff features.

Then, we quantify the contribution of *X*_*j*_ to (*Y, δ*) by the following two measures:

1. 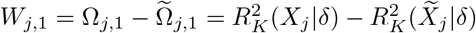.
2. 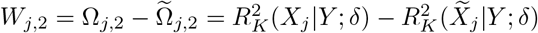.

Given sample data 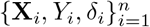, we estimate *W*_*j*,1_ by 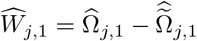, where 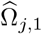 and 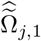 are calculated using (6) Similarly, we estimate *W*_*j*,2_ by 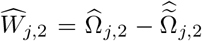 using (7). Intuitively, a large value of either 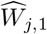 or 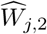 indicates the significance of *X*_*j*_ as *X*_*j*_ outperforms 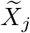. On the other hand, it is expected that irrelevant variables behave similarly to their knockoff counterparts, resulting in small sample utilities that bounce around 0 as shown in the following proposition.

#### Proposition 3

*Let* 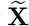 *be an exact knockoff copy of* **X**. *Then, for j /*∈ *𝒜*,

1. *W*_*j*,1_ = *W*_*j*,2_ = 0;
2. *Conditioning on* 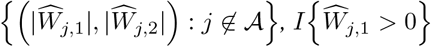 *and* 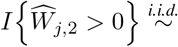 *Bernoulli(0*.*5)*,

*where I{·} is the indicator function*.

For fixed thresholds *t*_1_, *t*_2_ *>* 0, the false discovery proportion is

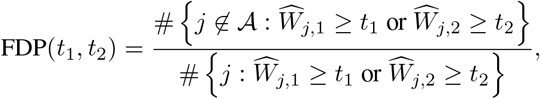

and FDR(*t*_1_, *t*_2_) = *E*[FDP(*t*_1_, *t*_2_)]. Note, from Proposition 3,

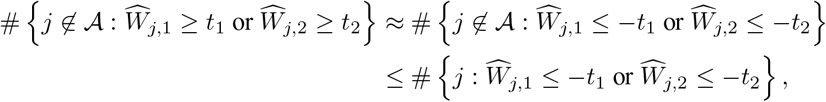

which leads to a conservative estimator of FDP:

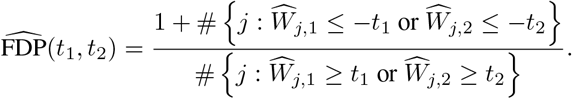

The proof of Proposition 3 appear in Appendix 6.4. The offset of 1 in the numerator, yielding a slightly more

conservative{_es(timator, is neces)sary both theoretic}ally and empirically to control the FDR. Define 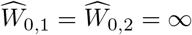 and let 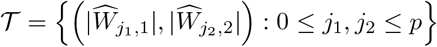. Then (*T, ::<*) is a partially ordered set, where (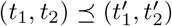 if 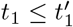 and 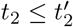, for (*t*_1_, *t*_2_), 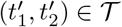. To control FDR at a pre-specified level *α*, we choose the thresholds *T*_*α*,1_ and *T*_*α*,2_ as

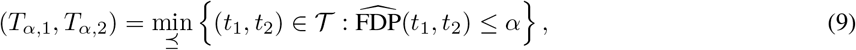

where min_*-<*_ represents the minimal element of the set with respect to *::<*. Then, the selected active set is given by

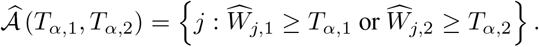

Note that there can be more than one minimal element in *T* ; so, the choice of (*T*_*α*,1_, *T*_*α*,2_) may not be unique, leading to different estimates of the active set. In practice, one can choose the minimal element that yields the largest average utility of the selected features, 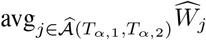, where 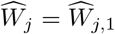 if *X*_*j*_ is ranked higher based on the marginal statistic than the conditional statistic and 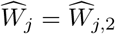 vice versa. This approach works well in our simulation studies (see, Section 3).

Although this dual selection procedure controls FDR, it is not readily applicable to ultrahigh dimensional data because constructing knockoff features becomes computationally intractable for large *p*. However, feature screening and knockoff-based selection naturally complement each other under ultrahigh dimensionality: one can perform screening to reduce *p*, and then apply the knockoff technique to further control FDR [3, 33]. We elaborate this adaption in the next subsection and show that sure screening is still attainable with FDR under control.

### 2.5 Refined Screening with FDR Control

Consider splitting *n* observations into two disjoint groups of size *n*_1_ and *n*_2_ = *n* − *n*_1_, denoted as 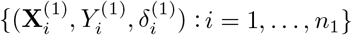 and 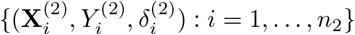. We follow the next two steps:

1. We start with conducting the screening procedure as described in 2.3 using 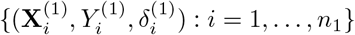 to select a small index subset of potentially relevant features 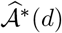 for *d < n*_2_.
2. Next, we run the knockoff procedure as described in 2.4 on the remaining data 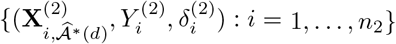, ignoring features that were not selected in the screening step. Specifically, we first obtain the knockoff matrix 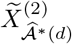 for the original design matrix 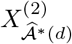 on the remaining data. Then, we compute 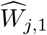 and 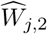 for 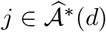. For a pre-specified FDR level *α*, we ultimately select

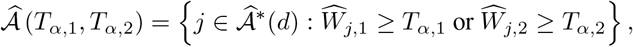

where *T*_*α*,1_ and *T*_*α*,2_ are determined by solving (9).

Hereafter, we refer to this screening-and-knockoff procedure as *α*-controlled *k*ernel-based *i*ndependence *d*ual *s*reening (*α*-KIDS for short). It is critical that the two steps of *α*-KIDS are performed on distinct data. The following procedure would not control the FDR: we perform the screening step using the full data set to select 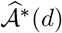 and run the knockoff procedure on the dimension-reduced data 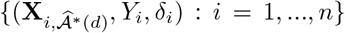. The problem is that 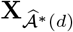 can be viewed as a function of (**X**, *Y, δ*) because 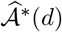 is selected using all data. As a result, there is no guarantee that 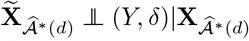, even if 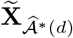 is constructed without using (*Y, δ*). The loss of FDR control is not merely theoretical; an unimportant feature *X*_*j*_ that is kept by the screening step is generally more likely to appear as a false positive when running the knockoff filter, leading to a much higher FDR [3]. With the data splitting mechanism, as long as the screening step correctly identifies all the relevant features (which happens asymptotically almost surely as shown in Theorem 2.3), the knockoff step will control the FDR as desired. Moreover, the sure screening property is inherited. That is, the *α*-KIDS procedure achieves a balancing trade-off between type I and type II errors. This appealing property is justified in Theorem 2.5 below.

Denote 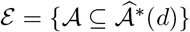 the event that all the important features are selected in the screening step. We further require that the true signal cannot be too weak to be captured by the knockoff filter:

lvbel=(C0’),vtemsep=0pt There exist *c*_5_, *c*_6_ *>* 0 and *γ*_5_, *γ*_6_ ∈ [0, 1*/*2), such that

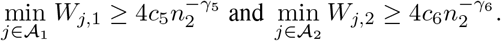

[FDR-Controlled Sure Screening] For any *α* ∈ (0, 1), we have

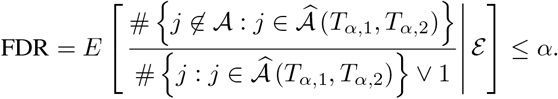

Furthermore, under condition v,

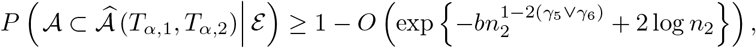

for *α* ≥ 1*/*|*A*|, where *b >* 0 is a constant.

The proof of Theorem 2.5 appear in Appendix 6.5. Although data splitting is a straightforward approach to handle ultrahigh-dimensionality, there is certainly a loss of power since each step only uses part of the data. One solution to the issue is to smartly recycle the data used in the screening step to raise power while retaining the FDR control property for the knockoff procedure [3]. We modify the *α*-KIDS procedure as follows:

1. The screening step remains the same. Use 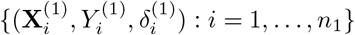 to select a small index subset of potentially relevant features 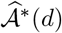 for *d < n*_2_.
2. For the knockoff step, we still start with obtaining the knockoff matrix 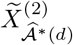 for the original design matrix 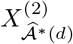 on the remaining data. Then we concatenate the original design matrix on the first *n*_1_ observations with the knockoff matrix on the next *n*_2_ observations as

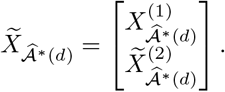

Now, we calculate 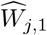 and 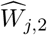 using the full data 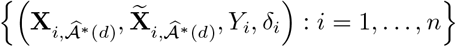 for 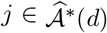, where 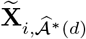 is the *i*th row of the knockoff matrix 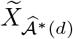. For a pre-specified FDR level *α*, we ultimately select

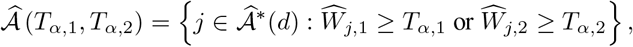

where *T*_*α*,1_ and *T*_*α*,2_ are determined by solving (9).

Here, we follow the convention to treat 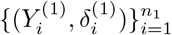 as fixed when creating 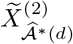 in the knockoff step [3]. In other words, although 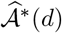 was selected using the first portion of the data, we think of 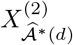 as being independent of 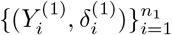. As a result, 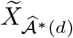 gives legitimate knockoff features and the procedure controls the directional FDR. On the other hand, there is an inherent gain of power compared to the data splitting approach as the first *n*_1_ observations weigh in. If a feature *X*_*j*_ is important, the first portion of data will contribute to large *R*^2^ or partial *R*^2^ values for both *X*_*j*_ and 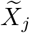 since 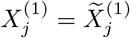 by design, and the second portion of data will help separate *X*_*j*_ from its knockoff counterpart, resulting in a positive value of *W*_*j*,1_ or *W*_*j*,2_.

## 3 Simulation Studies

In this section, We evaluate the performance of our method on simulated ultra-high dimensional datasets, and make comparisons with several other competing methods, including censored rank independence screening (CRIS) [40], integrated powered density (IPOD) [26], and robust censored distance correlation screening (RCDCS) [7]. Our method is conducted with the Gaussian kernel being the reproducing kernel as well as the smoothing kernel for density estimation. The bandwidths of the two Gaussian kernels are set to heuristic median pairwise distance [22] and 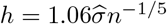, where *n* is the sample size and 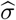 is the sample standard deviation [39]. We generate correlated features **X** from *N*_*p*_(**0**, Σ) with *p* = 5, 000 and Σ having a first-order autoregressive (AR) structure, and consider a variety of survival models under independent or dependent censoring. The design of correlated features is to mimic the phenomenon that features tend to be correlated, even purely by chance, in ultrahigh dimensional space [15], which makes it more challenging to distinguish truly important features from spurious ones and achieve exact feature selection. We report the following results based on 200 replicates:

- the *τ* ^*th*^ quantiles of the minimum model size (MMS), denoted as *M*_*τ*_, that includes all active features for the screening methods, where the MMS for KIDS is defined as min*{M*_1_ +*M*_2_*}* such that 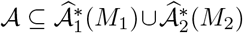;
- the proportion of selecting a certain active predictor *X*_*j*_, denoted as *P*_*j*_, and the proportion of including all active predictors, denoted as *P*_*A*_, for all the screening methods and *α*-KIDS;
- the average model size (AMS) determined by *α*-KIDS;
- and empirical FDR (EFDR) for *α*-KIDS.

### Example 1

In this example, we evaluate the efficacy of KIDS in comparison to the other screening methods. Let **X** ∼ *N*_*p*_(**0**, Σ), where Σ = *AR*(0.5). Given **X**, the true survival time is generated from the following accelerated failure time (AFT) model and proportional hazard (PH) model:

**1. Model 1:** 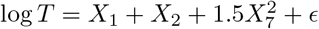, where *E* ∼ *N* (0, 1) independently;

**2. Model 2:** 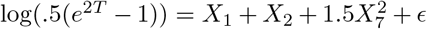, where *E* follows the standard extreme value distribution independently, which corresponds to a PH model [27].

For each model, the survival time is subject to two censoring mechanisms:

1. independent censoring time *C* generated from uniform distribution on [0, *c*_0_];
2. dependent censoring time *C* generated from exponential distribution with mean 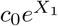, where, the constant *c*_0_ is chosen to achieve 30% or 50% censoring rate (CR).

The results are summarized in Table 1 for *n* = 200 and *d* = [*n/* log *n*] = 38. In all scenarios, KIDS outperforms the other methods with higher selection proportions, and minimum model sizes closer to the truth, i.e., | *𝒜*| = 3. The three competitors are not as robust to heavy censoring or dependent censoring as KIDS. In addition, CRIS barely detects the feature (*X*_7_) that is non-linearly related to the endpoint. In the Supplements, we further consider a linear design with varying signal strength (Example 3), and a more complex nonlinear design (Example 4) for both AFT- and PH-type of models under varying censoring mechanisms. Once again, our method performs consistently well compared to the other methods.

**Table 1:**
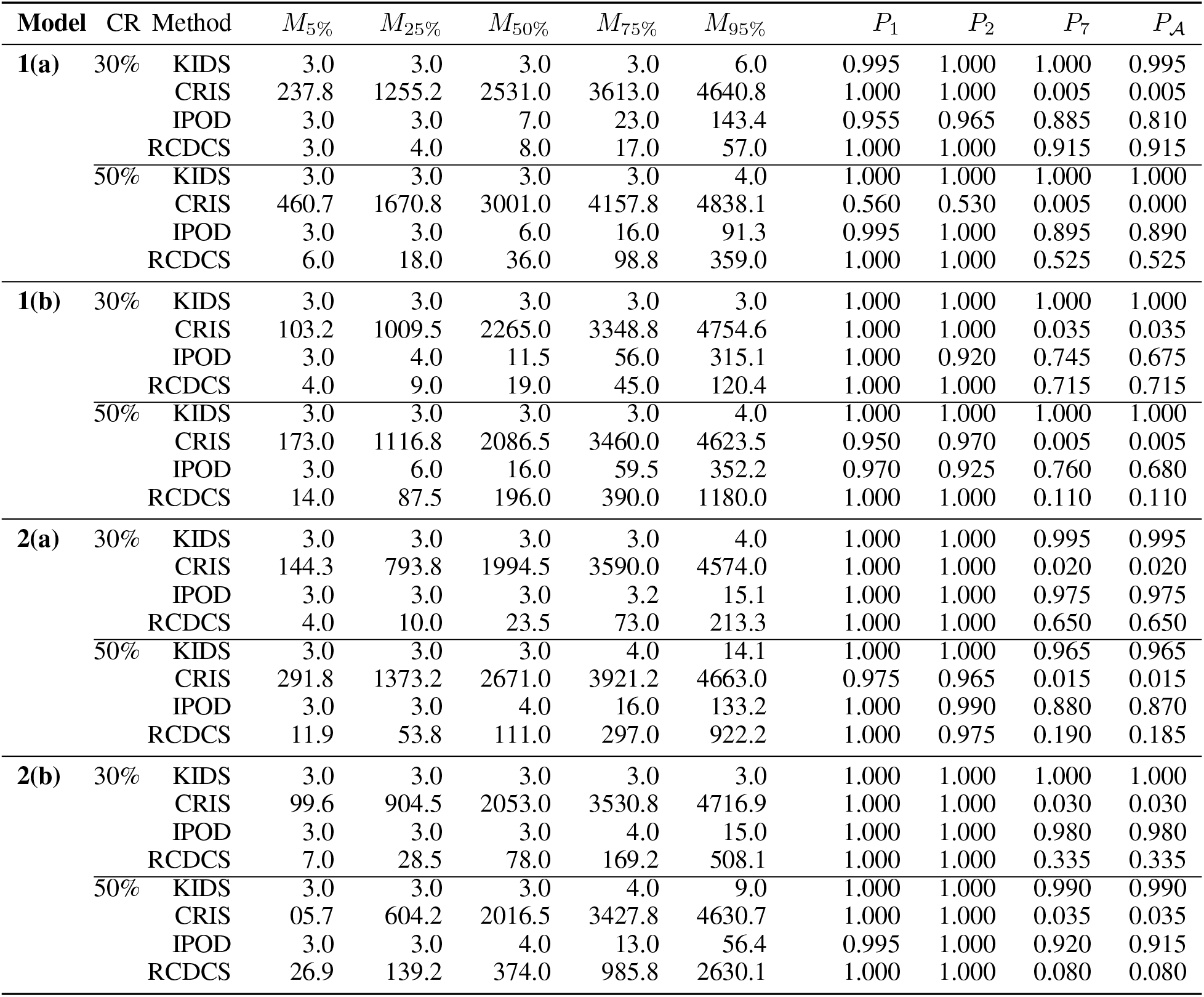
Quantiles of MMS (*M*_*τ*_) and selection proportions (*P*_*j*_’s and *P*_*A*_) for models in **Example 1** based on 200 replicates with *n* = 200, *p* = 5000 and *d* = [*n/* log *n*] = 38.

### Example 2

This example is to verify Theorem 2.5 for the *α*-KIDS procedure. Similar to Example 1, we generate **X** from *N* (**0**, Σ) with Σ = *AR*(0.3) and simulate the true survival time from the following two models:

**1. Model 3:** log *T* = *µ*(**X**) + *E*, where 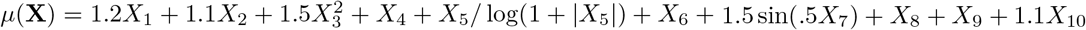 and *E* ∼ *N* (0, 1), independently;

**2. Model 4:** log(.5(*e*^2*T*^ − 1)) = *µ*(**X**) + *E*, where *µ*(**X**) is the same as in Model 3 and *E* follows the standard extreme value distribution, independently.

The censoring time is simulated from:

(a) uniform distribution on [0, *c*_0_];

(b) exponential distribution with mean 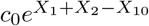,

where, the constant *c*_0_ is chosen to yield 30% or 50% CR.

We set *n* = 2, 000, *n*_1_ = 500, *n*_2_ = 1, 500, *d* = 100 and vary the nominal level *α* from .1 to .3[3, 33]. We report the overall selection proportion 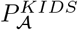 for the screening step, whereas, for the knockoff step, we report *P*_*j*_’s, *P* _*𝒜*_, AMS and EFDR given 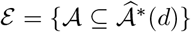. The results are summarized in Table 2. Despite the models involve linear and nonlinear terms of correlated features, the *α*-KIDS procedure in general controls the FDR at the desired level fairly well and inherits the sure screening property across different censoring settings. Note, at *α* = .1, the procedure has to precisely identify *𝒜* (in theory) to control the FDR and maintain power simultaneously because the FDP is exactly .1 with | *𝒜*| = 10 – a challenging borderline scenario. If *α <* .1, with high probability, the procedure ends up with an empty set.

**Table 2:**
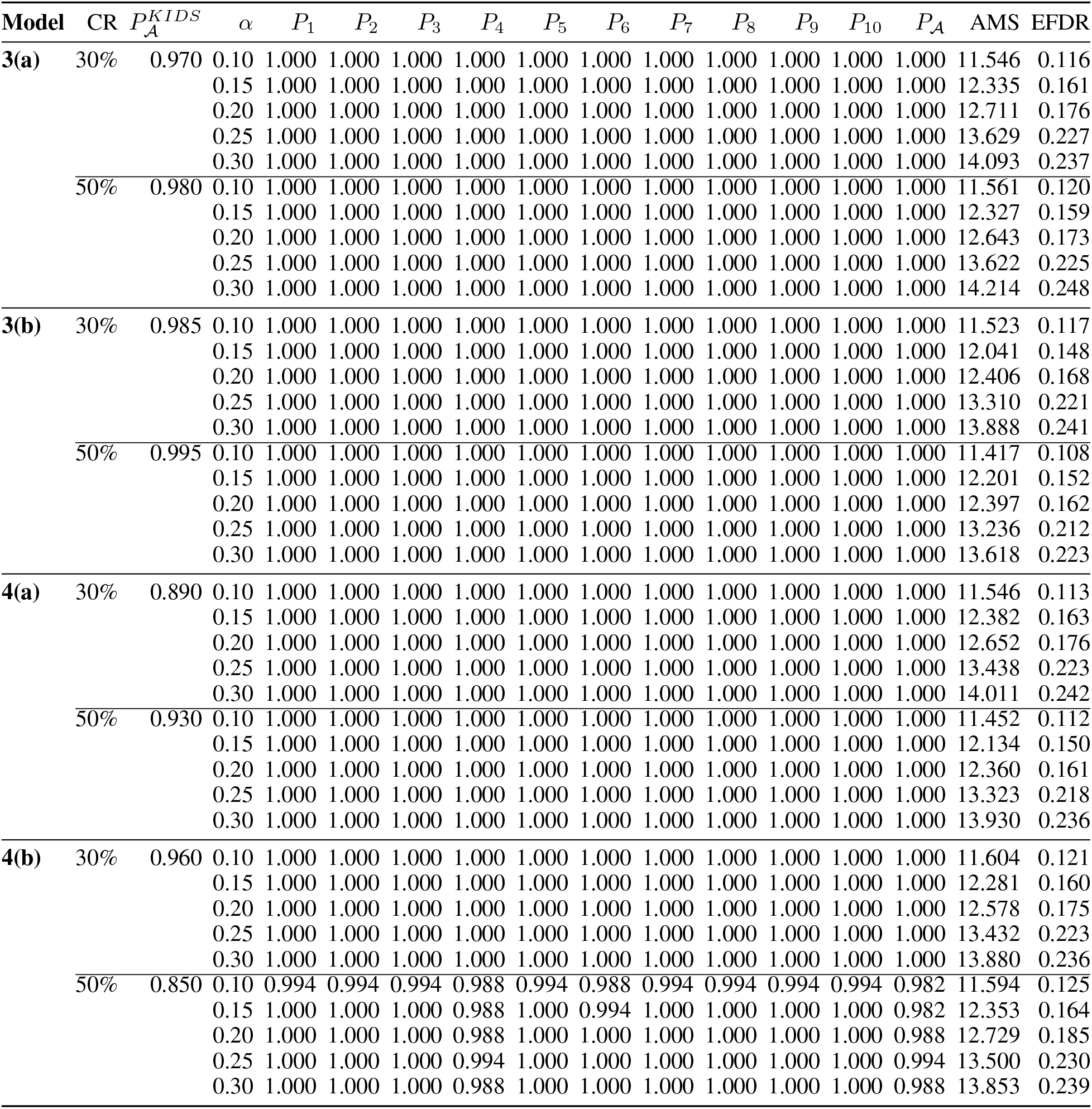
Selection proportions (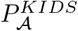, *P*_*j*_’s and *P*_*A*_), AMS and EFDR for models in **Example 2** based on 200 replicates with *n*_1_ = 500, *n*_2_ = 1500, *p* = 5000 and *d* = 100.

The choices of *n*_1_*/n*_2_ ratio and the model size *d* for the screening step in our setting appear to give a favorable balance between finding a sufficiently good screened set at the first stage, and retaining a large enough sample size for powerful inference in the second stage. In practice, we also suggest a split with *n*_2_ *> n*_1_ to allow more information for accurate selection via knockoff. On the other hand, *d* cannot be too small to ensure the coverage of *𝒜* for the screening step and to provide adequate amount of noise as reference to control FDR for the knockoff step, while the computational cost for knockoff may prevent us from choosing an arbitrarily large *d*. Whether we can determine theoretically the optimal split and model size for making the most discoveries is worthy of future research.

## 4 Application: Head And Neck Cancer Data

We investigated the head and neck squamous cell carcinoma (HNSCC) cohort in the Cancer Genome Atlas (TCGA) network. Upper quartile normalized RSEM TPM mRNA expression values for 518 primary-solid tumor samples with matched clinical information were obtained using the R package curatedTCGAData. Genes with low expression, as indicated by a zero interquartile range, were excluded from the analysis. The remaining genes were log-transformed. The endpoint of interest in our study was the number of days to death, which was subject to right censoring either due to loss of follow-up or no event occurrence until the end of the study. The observed survival time ranged between 2 to 6417 days with a median of 649.5 days and with 57.53% censoring rate. For external validation of our findings, gene expressions and clinical data for 253 HNSCC primary tumor samples were acquired from the Gene Expression Omnibus (GEO) database (accession number GSE65858 [47]). After preliminary data processing, a total of 15,887 genes commonly available in the TCGA and the GSE65858 datasets, along with important clinical covariates (as summarized in Table 8 in the Web Supplement), were included for our analysis. The R package DSFDRC available in GitHub (https://github.com/urmiaf/DSFDRC) implements the methodology.

### 4.1 Model Selection

The TCGA samples were partitioned into training and testing subgroups using a 4:1 ratio. The training cohort consisted of 414 samples, while the testing cohort had 104 samples, and both cohorts shared the same censoring proportion. We employed various competing methods on the training data to identify prognostic gene signatures for HNSCC, and subsequently evaluated their performance using the testing samples. The models were as follows:

- *α*-KIDS, with specific parameters set to *n*_1_ = 200, *d* = 100 and *α* = 0.1, followed by a Cox proportional hazard model built on the selected genes;
- KIDS/CRIS/IPOD/RCDCS to pre-select *d* = [104*/* log(104)] = 22 candidate genes, followed by a penalized Cox model (CoxNet) applied on the dimension-reduced data for further gene selection and prognostic modeling;
- *α*-KIDS followed by a Cox gradient boosting machine (CoxGBM [18]) built on the selected genes;
- KIDS/CRIS/IPOD/RCDCS to pre-select *d* = 22 candidate genes, followed by the double-slicing assisted procedure (DS [9]) for further gene selection and finally a prognostic CoxGBM applied on the dimension-reduced data after the screening and selection steps.

To ensure a fair comparison between *α*-KIDS (which performs both screening and selection), and the screening-only procedures (KIDS, CRIS, IPOD, and RCDCS), we augmented the screening procedures with more precise selection techniques, namely CoxNet, a model-based approach, and DS, a model-free method. The DS procedure identifies low-dimensional sparse linear combinations of features Γ^*T*^ **X**, such that (*Y, δ*) **⊥ X**|Γ^*T*^ **X**, where Γ is a *p × q* matrix with *q* (the number of linear combinations) usually being much smaller than *p*. It achieves simultaneous feature selection through regularization without assuming any parametric distribution of (*Y, δ*) or linear relation between (*Y, δ*) and Γ^*T*^ **X**. For the purpose of gene selection, we only leveraged the ability of DS to extract relevant genes rather than utilizing the linear combinations it produced. Both linear Cox model and nonlinear CoxGBM were used to construct prognostic signatures based on the selected genes. Optimal tuning parameters for CoxNet, DS, and CoxGBM were determined through cross-validation.

A patient’s gene signature loading was calculated as the linear predictor for the fitted Cox/CoxNet model, or the link function value of the fitted CoxGBM model, which can also be viewed as a risk score. Subsequently, patients were classified into high-risk and low-risk groups, using the median risk score of the training cohort as the cutoff. The log-rank tests were conducted to compare the survival functions of the two risk groups and the p-values are reported in Table 3. Furthermore, we assessed the gene signatures by the time-dependent dynamic receiver operating characteristic (ROC) curves [24] at 1, 3 and 5 years. The corresponding area under curve (AUC) values are summarized in Table (3). According to the log-rank tests and the ROC curves, relevant genes selected by *α*-KIDS and KIDS led to more informative prognostic signatures for HNSCC in terms of risk stratification and survival prediction. Notably, the *α*-KIDS+CoxGBM model demonstrated the most favorable overall performance on the testing samples. We then proceeded to refit the model to the entire TCGA dataset, and validated the resulting gene signature with the external data.

**Table 3:**
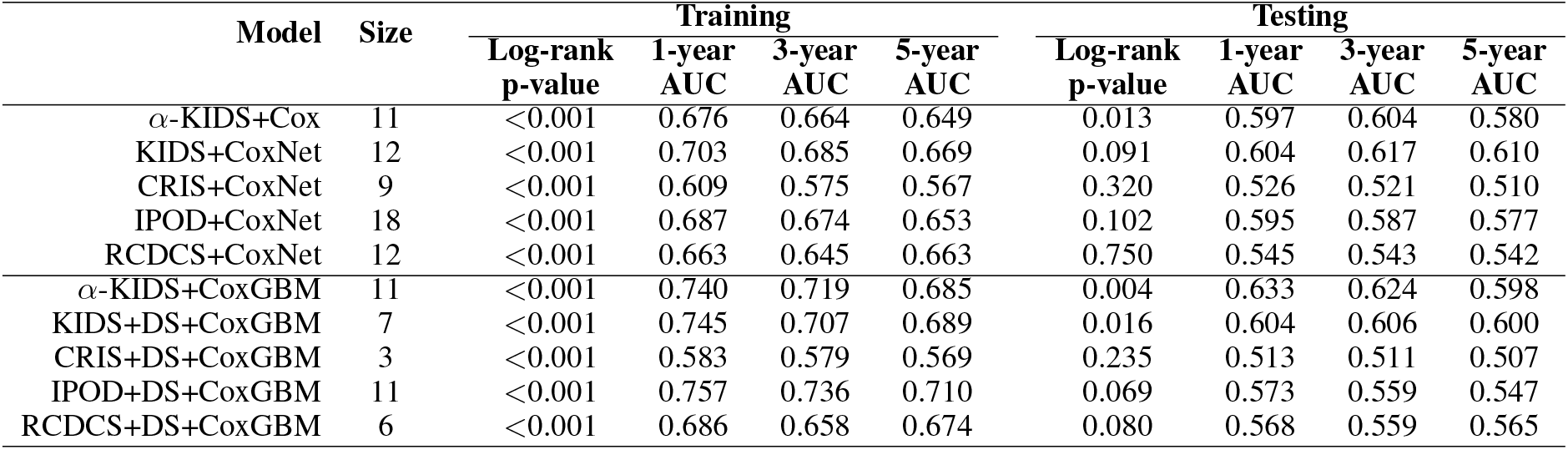
Gene signature sizes (number of genes in a signature), p-values for the log-rank tests on the risk stratification of the TCGA samples, and AUCs for 1-, 3-, 5-year ROC curves of the risk scores across competing methods.

### 4.2 10-gene Signature and External Validation

The *α*-KIDS procedure was applied to the full TCGA dataset, resulting in the selection of 10 genes: OLR1, SPOCK1, DDX19A, FADS3, P2RX6, C9ORF4, C15ORF21, TMED6, TFB2M and C22ORF15. A 10-gene prognostic signature was constructed using CoxGBM subsequently. The GEO platform data was used to validate the effectiveness of the gene signature. Patients were classified as having a high-risk gene signature or a low-risk gene signature on the basis of the link function values, with the median score of the TCGA samples as the threshold. Patients with a high-risk 10-gene signature exhibited significantly lower median survival compared to those with a low-risk gene signature in both the TCGA cohort (727 days vs. 2717 days) and the validation cohort (1068 days vs. 1962 days), as supported by the Kaplan-Meier curves and the log-rank test p-values (Figure 1). An interesting finding is that patients with HPV infection were associated with lower risk scores in both cohorts (p-values *<* 0.001 based on two-sample t-tests). This aligns with previous reports that patients with HPV-positive cancers generally experience better prognoses than those with HPV-negative cancers, particularly for tumors arising in the oropharynx [29]. Therefore, the gene signature may offer insights into the underlying molecular mechanisms of the HPV heterogeneity.

**Figure 1:**
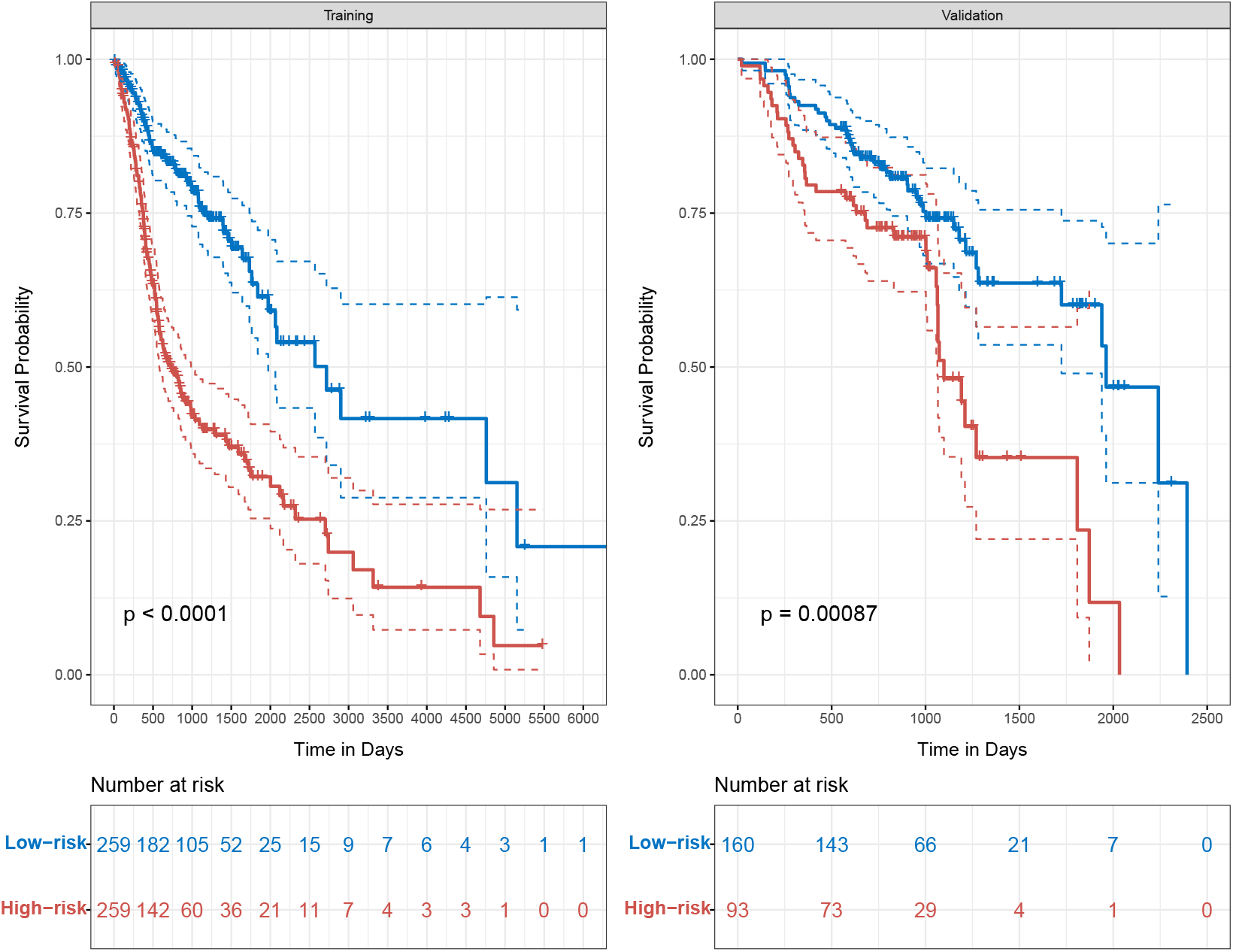
Kaplan–Meier estimates of overall survival (solid) with 95% confidence interval (dash) and log-rank test p-values for risk stratification of training (TCGA) and validation (GEO) samples according to the 10-gene signature identified by the *α*-KIDS+CoxGBM model.

Additionally, multivariate Cox proportion-hazards regression analysis was used to evaluate independent prognostic factors associated with survival, and the 10-gene signature, age, sex, tumor stage, HPV status, alcohol history and smoking history were used as covariates. The fitted models are summarized in Table 4. For the TCGA cohort, the 10-gene signature was a strong predictor with an hazard ratio of 7.62 (p-value *<* 0.001), after adjusting for other clinical covariates. There was a 2% increase in the expected hazard relative to a one year increase in age (p-value *<* 0.001). Patients with IV-stage cancer experienced a remarkable 98% increase in hazard (p-value *<* 0.001) compared to those in early stages (I/II). Similar results were observed in the validation cohort, indicating the potential clinical utility of the 10-gene signature in enhancing prognostic assessments and guiding personalized treatment decisions for HNSCC patients beyond conventional phenotype-based predictors.

**Table 4:** Multivariate Cox regression analysis based on the 10-gene signature and other clinical covariates. CI denotes confidence interval.

| Variable | Training (TCGA) |  | Validation (GEO) |  |
| --- | --- | --- | --- | --- |
|  | Hazard Ratio (95% CI) | P-value | Hazard Ratio (95% CI) | P-value |
| 10-gene signature | 7.62 (4.59, 12.66) | <0.001 | 3.39 (1.69, 6.79) | <0.001 |
| Age | 1.02 (1.01, 1.04) | <0.001 | 1.03 (1.00, 1.05) | 0.030 |
| Gender |  |  |  |  |
| Male | 1.00 (0.70, 1.43) | 0.989 | 0.92 (0.50, 1.67) | 0.773 |
| Female | - | - | - | - |
| Tumor stage |  |  |  |  |
| III | 1.55 (0.92, 2.60) | 0.099 | 1.07 (0.32, 3.57) | 0.915 |
| IV | 1.98 (1.32, 2.98) | <0.001 | 5.34 (2.29, 12.47) | <0.001 |
| I/II | - | - | - | - |
| HPV status |  |  |  |  |
| Positive | 1.53 (0.96, 2.47) | 0.353 | 0.51 (0.29, 0.91) | 0.021 |
| Negative | - | - | - | - |
| Alcohol history |  |  |  |  |
| Yes | 0.86 (0.61, 1.19) | 0.915 | 1.19 (0.57, 2.50) | 0.639 |
| No | - | - | - | - |
| Smoking history |  |  |  |  |
| Yes | 1.02 (0.70, 1.50) | 0.854 | 0.99 (0.52, 1.90) | 0.977 |
| No | - | - | - | - |

### 4.3 Integrated Clinicogenomics Modeling

Results from the above analysis motivated us to consider a CoxGBM combining the 10 genes selected by *α*-KIDS and important clinical covariates, namely age, sex, tumor stage, HPV status, alcohol history and smoking history. The composite model was compared against two other CoxGBM models: the one based solely on the 10 selected genes (which was used to discover the 10-gene signature in the previous subsection) and the other based solely on the clinical covariates. Again, the TCGA cohort was utilized as the training data and the GEO platform data served as the external validation data. Differences in survival between the high-risk group and the low-risk group were analyzed with the log-rank test. ROC curves and associated AUCs were calculated to assess time-dependent predictive performance of the three models. The results, as summarized in Table 5, revealed that the model integrating both clinical and genetic information had improved prognostic accuracy over the other two models.

**Table 5:** P-values for the log-rank tests on the risk stratification, and AUCs for 1-, 3-, 5-year ROC curves of the risk scores across three competing CoxGBMs.

| Variables | Model size | Training |  |  |  | Validation |  |  |  |
| --- | --- | --- | --- | --- | --- | --- | --- | --- | --- |
|  |  | Log-rank p-value | 1-year AUC | 3-year AUC | 5-year AUC | Log-rank p-value | 1-year AUC | 3-year AUC | 5-year AUC |
| Clinical-only | 6 | <0.001 | 0.626 | 0.606 | 0.611 | 0.012 | 0.642 | 0.605 | 0.613 |
| Genetic-only | 10 | <0.001 | 0.670 | 0.674 | 0.664 | <0.001 | 0.621 | 0.619 | 0.579 |
| Composite | 16 | <0.001 | 0.720 | 0.678 | 0.665 | <0.001 | 0.698 | 0.693 | 0.620 |

Finally, we highlight some biological implications of the genes selected by *α*-KIDS. OLR1 is a scavenger receptor for oxidized low-density lipoprotein (LDL) on endothelial cells and other cell types. OLR1 up-regulation in different tumors has evidenced its involvement in cancer onset, progression and metastasis, including HNSCC [36, 50]. High expression of FADS3, located at the cancer genomic hotspot 11q13 locus, has been reported to predict poor prognosis in HNSCC [41]. The oncogenic functions of SPOCK1, C15orf21, and TMED6 have also been investigated in several cancer cells [38, 17, 44, 11, 49].

## 5 Discussion

Large scale collaborative effort, such as TCGA, have allowed researchers with access to vast and curated data, enabling investigations into the underlying molecular mechanisms of HNSCC prognosis at various levels of complexity. A notable characteristic of such datasets is their ultra-high dimensionality, which places particular demands on the methods used to build prognostic models they must be able to handle data where the number of features far exceeds the number of observations. Moreover, in the context of survival analysis, how to handle censoring appropriately is paramount to avoid biased estimations and drawing incorrect conclusions, especially in the presence of heavy censoring. Feature screening emerges as a crucial step to efficiently reduce dimension before undertaking more accurate analyses. However, existing methods often impose explicit or implicit assumptions on censoring that are rather difficult to verify given the large number of features, creating impediments to their practical uses. Our proposed novel feature screening procedure quickly reduces irrelevant information under ultrahigh-dimensional right-censored settings, along with a unified selection procedure to control FDR. The proposed framework requires no pre-specification of the model structure and has the minimal assumption on the censoring mechanism. The flexibility is achieved by direct nonparametric learning of the survival outcome, without the need for intermediate estimation of survival probabilities. Our methodology is also readily generalizable for feature evaluation in other cancer types.

We remark that even if our assumption in equation 3 is not met, our procedure still serves the purpose of feature screening by identifying *𝒜* _(*T,C*)_, the active set for (*T, C*), jointly, which inherently contains *A*_*T*_. To further isolate the important features for *T* from the estimated active set, more precise feature selection methods [9] that are tailored for lower dimensional data, can be further applied. The FDR control step ensures that only the most informative features enter the downstream analyses to construct accurate prognostic models. Although our initial motivation was to address the challenges lying within the TCGA HNSCC dataset, the developed method is generally applicable to devise robust prognostic systems for new patient cohorts and other cancers. Along the line, future research should explore how to account for sample heterogeneity and integrate domain knowledge into the feature screening procedure, especially for HNSCC data encompassing samples from diverse sites and HPV subtypes. Additional avenues for future research include extending the methodology to more complex (cancer) endpoints, such as interval-censoring, and multistate models, etc. Our current exploration only considers patient mRNA genomic information. However, an integrative approach that analyzes and combines multiple -omics data, such as genomic, transcriptomic, and methylome data via identifying and validating a multi-omics signature may enhance HNSCC prognosis[37]. An extension of our current methodology for the multi-omics case, although non-trivial, is relevant.

## Data Availability

All data produced in the present study are available upon reasonable request to the authors

https://github.com/urmiaf/DSFDRC

## Supplementary Materials

## 6 Theorems and Proofs

### 6.1 Lemmas and Proofs

In this subsection, we first show some useful lemmas as preliminary results for Supplements 6.2 and 6.3.

#### Lemma 1

(Deviation bound for U-statistics, [25]). *Let g*(**U**_1_, …, **U**_*r*_) *be a kernel of a U-statistic U*_*n*_, *i*.*e*., 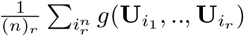, *where n > r*, 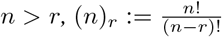 *and is taken over all r-tuples {i*_1_, …, *i*_*r*_*} drawn without replacement from {*1, …, *n}. If b*_1_ ≤ *g*(**U**_1_, .., **U**_*r*_) ≤ *b*_2_, *then for any E >* 0, *the following bound holds:*

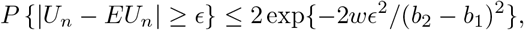

*where w* := [*n/r*], *the largest integer contained in n/r*.

This lemma gives a uniform bound for any U-statistic of arbitrary dimensional data, as long as the associated kernel is bounded. We repeatedly use this result to prove the next two lemmas.

#### Lemma 2

(Deviation bound for marginal utilities). *Under condition i, for any E* ∈ (0, 1),

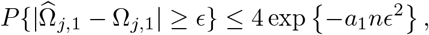

*where j* = 1, …, *p, and a*_1_ *>* 0 *is a constant*.

*Proof*. We aim to show the uniform consistency of the denominator and the numerator of Ω_*j*,1_ under regularity conditions respectively. Because the denominator of Ω_*j*,1_ has a similar form as the numerator, we deal with its numerator only below. Let

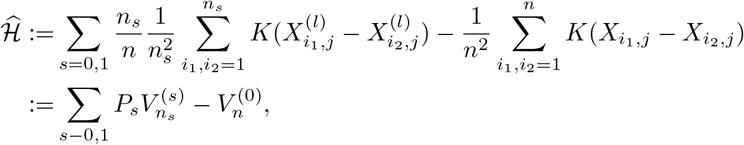

where 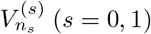 are V-statistics. Let 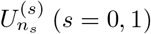 be corresponding U-statistics with 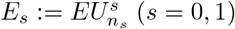. Under condition i, without loss of generality, we assume that the kernel *K* is bounded above by 1. Hence, 0 ≤ *E*_*l*_ ≤ 1 for *s* = 0, 1. Denote 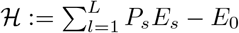, where *P*_*s*_ = *P* (*δ* = *s*). For any *E* ∈ (0, 1),

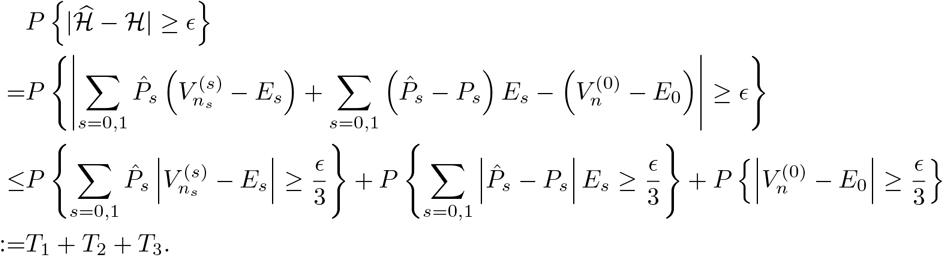

Let us consider *T*_1_ first.

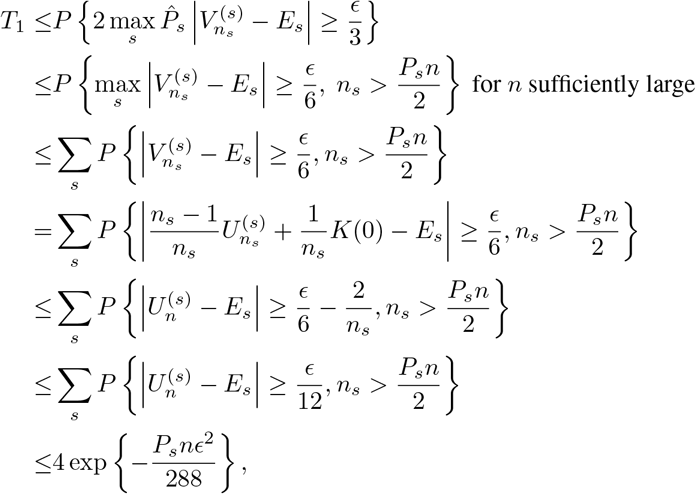

where the last inequality follows from Lemma 1. Also,

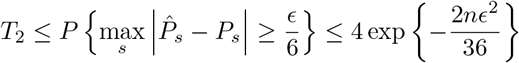

and 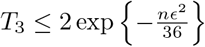. Combining *T, T* and *T*, we have

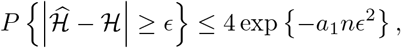

for some *a*_1_ *>* 0.

#### Lemma 3

(Deviation bound for conditional utilities). *Under conditions i-iv, for any E* ∈ (0, 1),

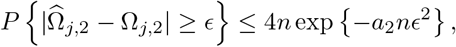

*where j* = 1, …, *p, and a*_2_ *>* 0 *is a constant*.

*Proof*. For a given *j* ∈ *{*1, …, *p}* and *s* ∈ *{*0, 1*}*, let 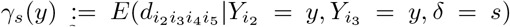, where 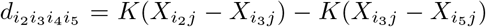, then 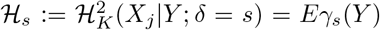. The kernel regression estimator

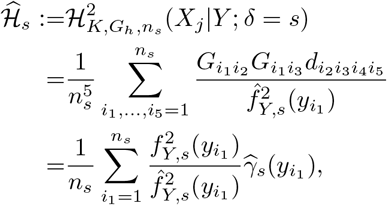

where 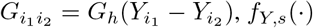 is the density function of *Y* given 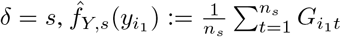 and

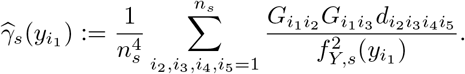

Without loss of generality, we assume that *f*_*Y,s*_(*y*) is bounded below by some *L >* 0 in condition iv. We first show some intermediate results. libel=(R0),leftmirgin=2 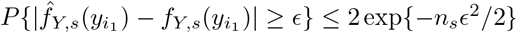 Note that 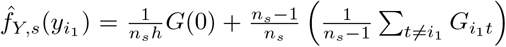 and 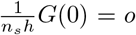 conditions ii and iii. Denote 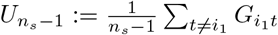. Then

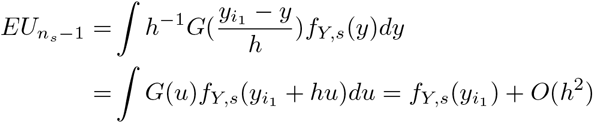

by Taylor expansion and conditions ii and iv. Hence,

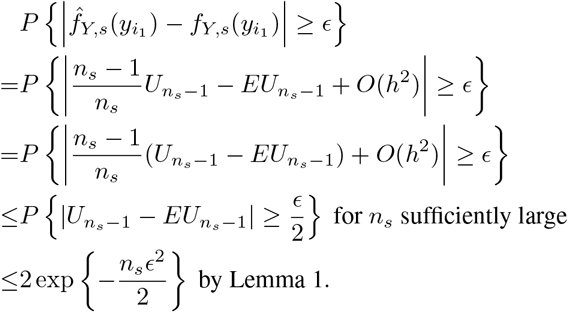

liibel=(R0),leftmiirgiin=2 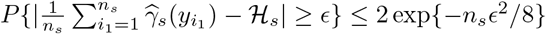.

Denote the corresponding U-statistic of 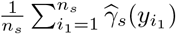 as 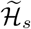, that is,

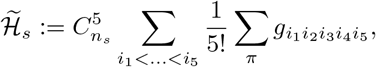

Where 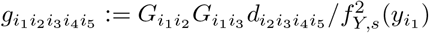 and Σ _π_ represents summation over the 5! permutations of (*i*_1_, …, *i*_5_). Under conditions ii and iii, 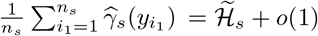. We will show in the next that 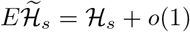 in two parts. Firstly,

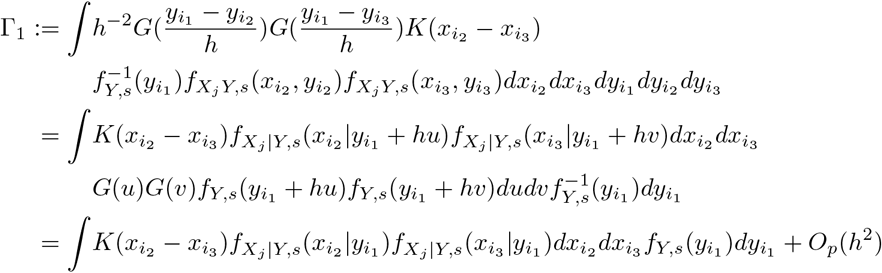

by Taylor expansion and conditions ii and iv. Similarly, we can show

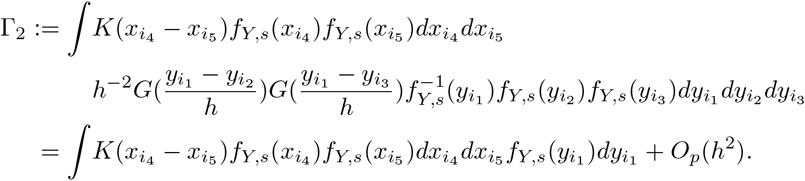

Therefore, 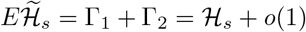. Then

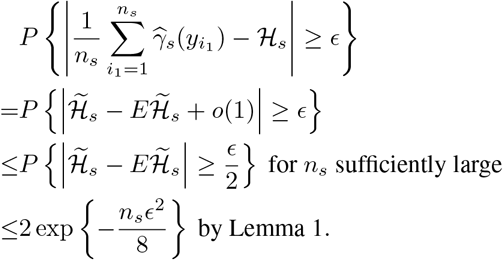

Now, for arbitrary ϵ ∈ (0, 1),

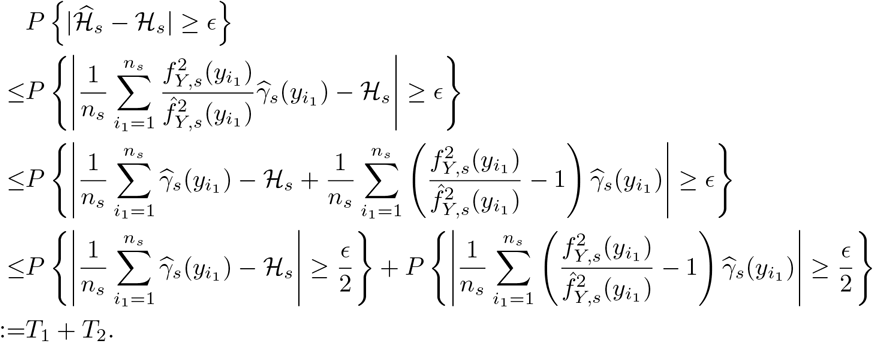

By (R2), *T*_1_ ≤ 2 exp*{*−*n*_*w*_*ϵ*^2^*/*32*}*. Moreover,

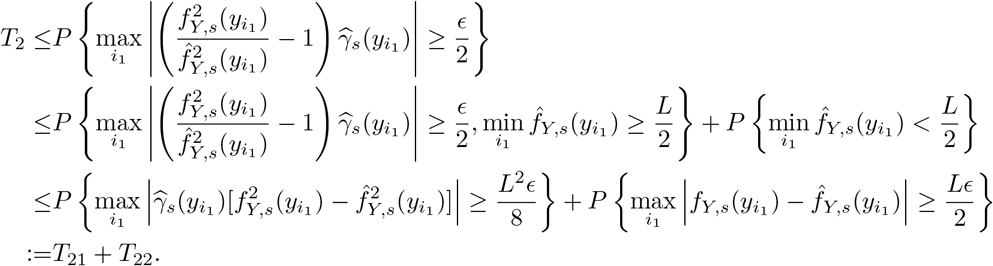

By (R1), 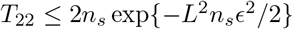. Let 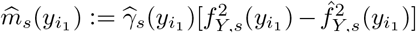 and 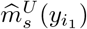 be the corresponding U-statistic. Similar to (R2), we can show that

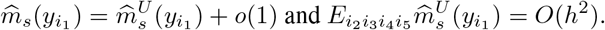

Hence, for *n*_*s*_ sufficiently large,

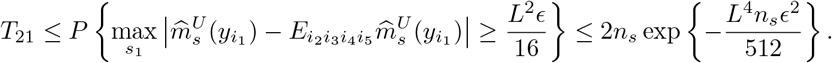

Finally, we have

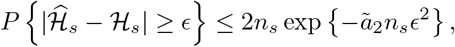

Where 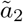 is some constant depending on *L*. Consequently,

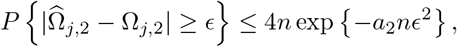

for some *a*_2_ *>* 0.

### 6.2 Proof of Theorem 2.3

*Proof*. Following from **Lemma 2** and **3**,

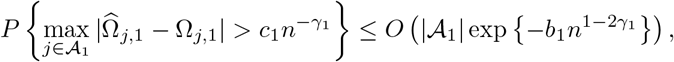

And

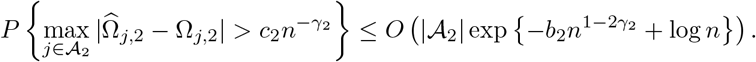

Under condition v, if 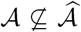, there must exist some *j* ∈ *𝒜*_1_ such that 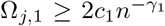 or some *j* ∈ *𝒜*_2_ such that 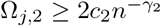 but 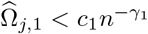 and 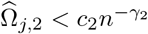. Therefore,

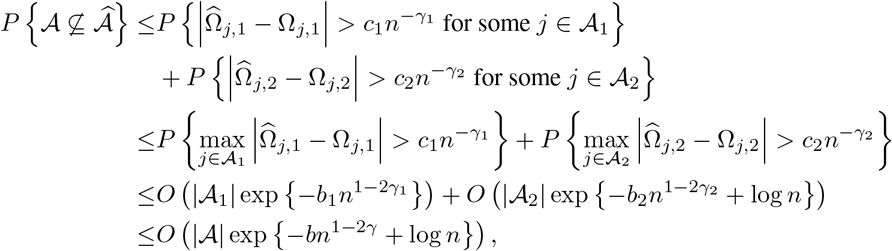

where *b* is a constant depending on *c* and *c*, and *γ* = max*{γ*_1_, *γ*_2_*}*. In other words,

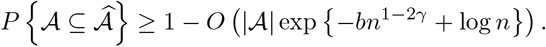

### 6.3 Proof of Theorem 2.3

*Proof*. By condition vi and **Lemma 2**,

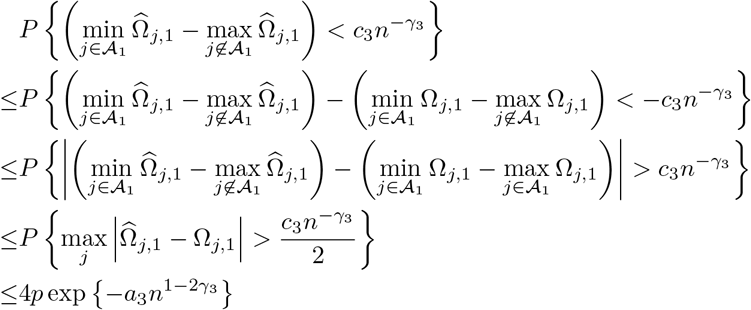

for some *a*_3_ *>* 0 depending on *c*_3_. Since 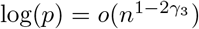, we have 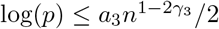 for *n* sufficiently large. For some *n*_0_ sufficiently large,

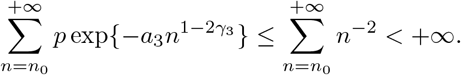

By Borel-Cantelli Lemma,

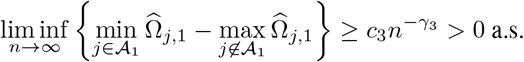

We can derive similarly that

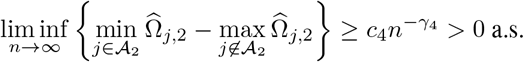

### 6.4 Proof of Proposition 3

*Proof*. For any *j*, let (**X**, 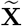)_(*j*)_ be the vector by swapping the entries *X*_*j*_ and 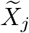 in (**X**, 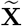); let (**x**, 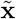)_(*j*)_ be the vector by swapping the entries *x*_*j*_ and 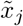 in (**x**, 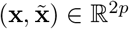) ∈ *ℝ*^2*p*^; and let **x**_−*j*_ denote the vector of **x** excluding *x*_*j*_. Let *f*_**U**|**V**_(**u**|**v**) denote the conditional distribution of **U** given **V** = **v**. For *j /*∈ *𝒜*_1_,

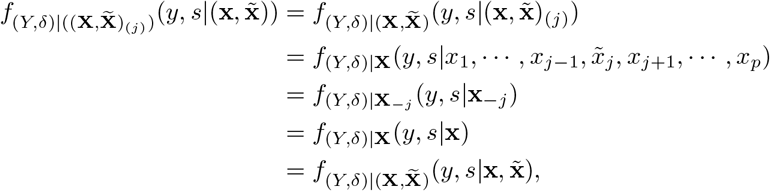

where the second and the last equations are due to (*Y, δ*)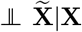, and the two equations in between follow from (*Y, δ*) **⊥** *X*_*j*_ |**X**_−*j*_ for *j /*∈ *𝒜*. That is,

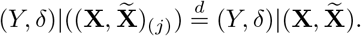

Since 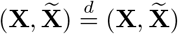 by the definition of knockoff copies, it follows that

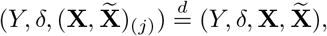

which implies that 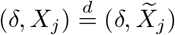 and 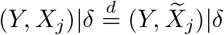. Therefore, *W*_*j*,1_ = *W*_*j*,2_ = 0.

In fact, we can show by repeating the above arguments that for any *𝒮* ⊂ *𝒜*^*c*^,

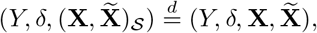

where (**X**, 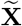)_*S*_ is the vector by swapping the entries *X*_*j*_ and 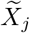 in (**X**, 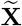) for all *j* ∈ *𝒮*. Let 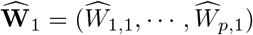 and let 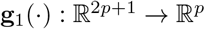 be a function such that 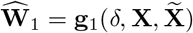. Define *E*_1_, *· · ·, E*_*p*_ such that *E*_*j*_ = 1 for *j* ∈ *A*_1_ and *E*_*j*_ is i.i.d. coin flip of *{*+1, −1*}* for *j /*∈ *𝒜*_1_. Consider *𝒮* = *{j* : *E*_*j*_ −1*}* ⊂ 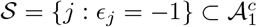, then

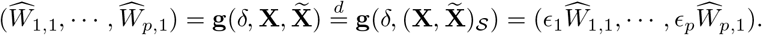

The statement for 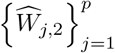 can be shown analogously.

### 6.5 Proof of Theorem 2.5

Throughout this proof, we restrict ourselves to the event 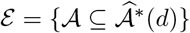. Let 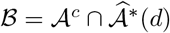. Denote by 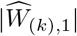 the *k*th largest absolute value of the marginal statistics 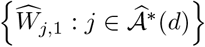, *k* = 1, …, *d*. Define 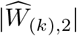 analogously for the conditional statistics. For ease of presentation, we further define 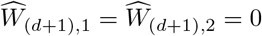. Then we have

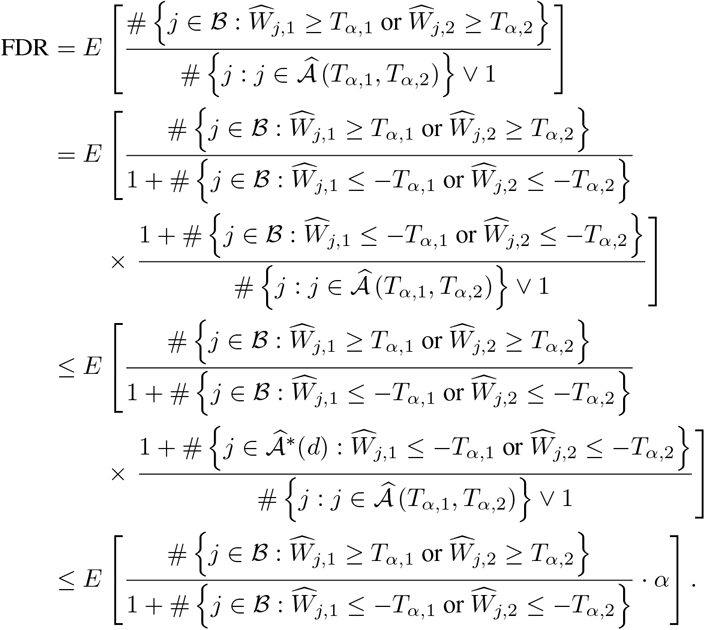

The first inequality holds since 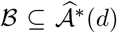 and the second inequality is due to the definition of (*T*_*α*,1_, *T*_*α*,2_) in (9). Consider a partially-ordered discrete time ocess

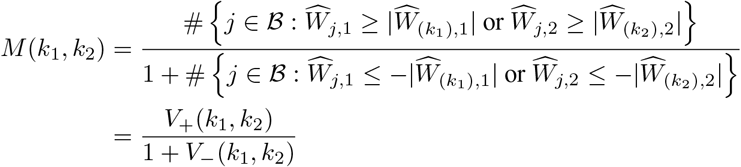

for 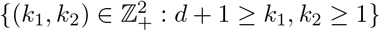, where

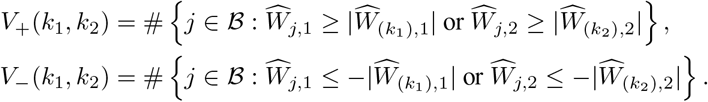

Let *F*(*k*_1_, *k*_2_) be the *σ*-field generated by knowing *{V*_*±*_(*j*_1_, *j*_2_) : *d* + 1 ≥ *j*_1_ ≥ *k*_1_, *d* + 1 ≥ *j*_2_ ≥ *k*_2_*}* as well as all the non-null statistics. The collection *{F* (*k*_1_, *k*_2_) : *k*_1_, *k*_2_ = *d* + 1, *d*, …, 1*}* of *σ*-fields is monotonic (thus a filtration) since 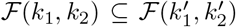 for 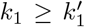 and 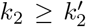. In the next we show that *{M* (*k*_1_, *k*_2_) : *k*_1_, *k*_2_ = *d* + 1, *d*, …, 1*}* is a supermartingale (running backward) with respect to *{F* (*k*_1_, *k*_2_) : *k*_1_, *k*_2_ = *d* + 1, *d*, …, 1*}*. In other words, *E* [*M* (*k*_1_ − 1, *k*_2_)|*F* (*k*_1_, *k*_2_)] ≤ *M* (*k*_1_, *k*_2_) and *E* [*M* (*k*_1_, *k*_2_ − 1)|*F* (*k*_1_, *k*_2_)] ≤ *M* (*k*_1_, *k*_2_), ∀*k*_1_, *k*_2_.

Suppose that 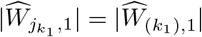 for 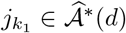. The filtration *F*(*k*_1_, *k*_2_) informs us about whether 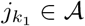 ∈ *𝒜* or not. On the one hand, if 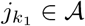 ∈ *A* or 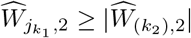, then *M* (*k*_1_ − 1, *k*_2_) = *M* (*k*_1_, *k*_2_). On the other hand, if 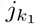 ∈ *ℬ* and 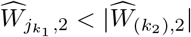, then

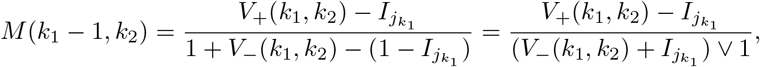

Where 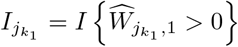. Since *I*_*j*_ 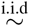 Bernoulli(0.5) for *j* ∈ *B* by Proposition 3, it follows that 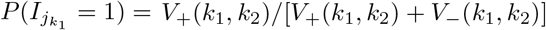 given ℱ (*k*_1_, *k*_2_). As a result,

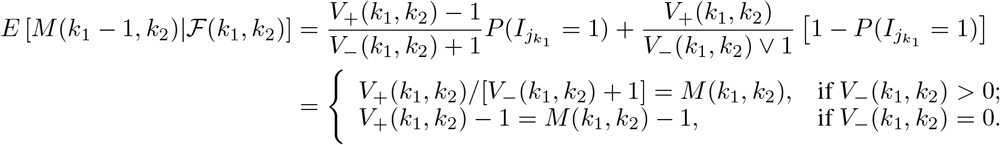

Therefore, *E* [*M* (*k*_1_ − 1, *k*_2_)| *ℱ* (*k*_1_, *k*_2_)] ≤ *M* (*k*_1_, *k*_2_). We can show that *E* [*M* (*k*_1_, *k*_2_ − 1)| *ℱ* (*k*_1_, *k*_2_)] ≤ *M* (*k*_1_, *k*_2_) in the similar vein.

In this process, (*T*_*α*,1_, *T*_*α*,2_) can be regarded as a stopping time with respect to the filtration ℱ (*k*_1_, *k*_2_) as 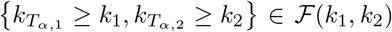 ∈ *ℱ* (*k*_1_, *k*_2_), where 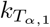 and 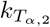 denote the indices such that 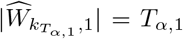 and 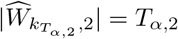. According to the optional sampling theorem [45] and Proposition 3, we deduce

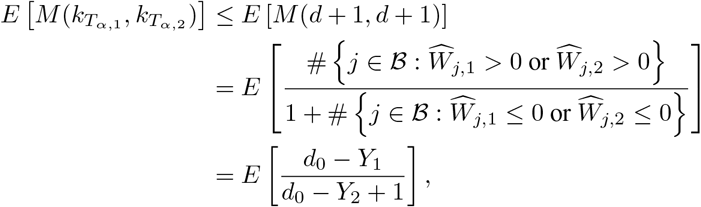

where *d*_0_ = |*ℬ*|, *Y*_1_ = # {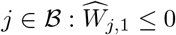 and 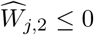}, *Y*_2_ = # {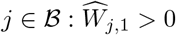 and 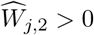}. Let *Y*_3_ = # {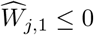 and 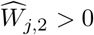} and *Y*_4_ = # {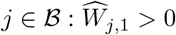 and 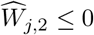}. Then (*Y*_1_, …, *Y*_4_) follow a multinomial distribution with equal event probabilities and 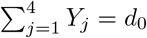. It follows that

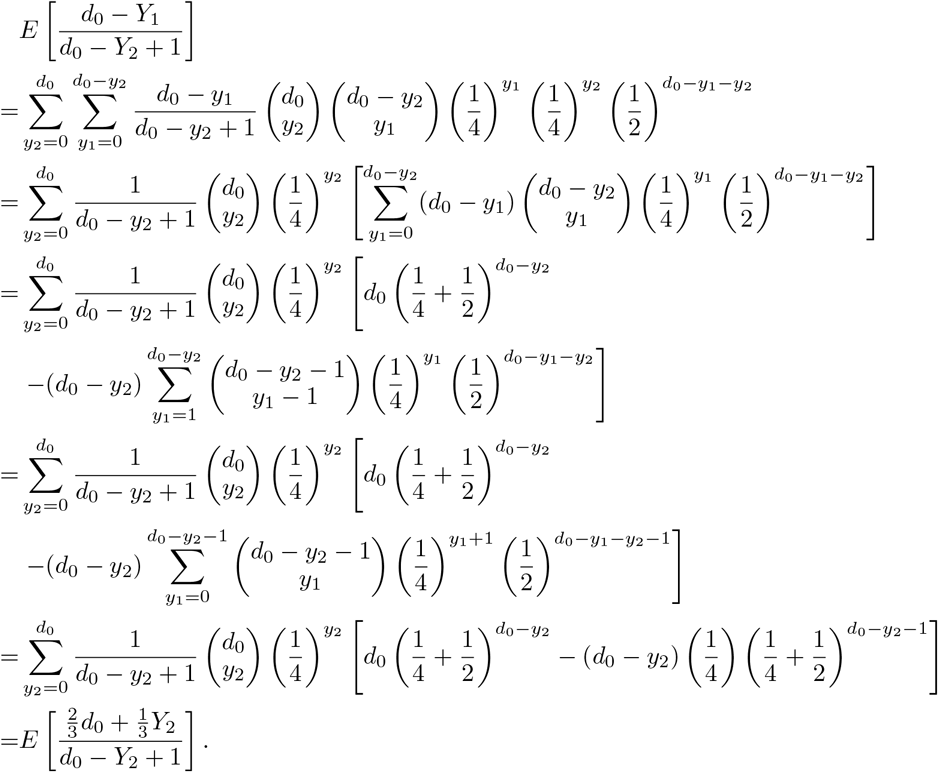

Since *Y*_2_ ∼ Binomial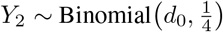 by Proposition 3, we have

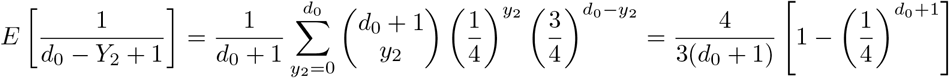

And

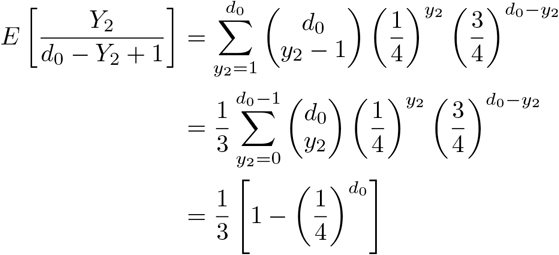

Therefore,

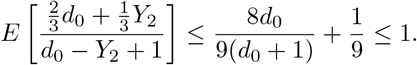

As a consequence, FDR≤ *α*.

In the next, we show the sure screening property. We can deduce from Lemma 2 that

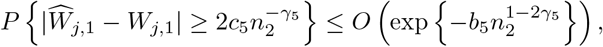

for some *b*_5_ *>* 0. Furthermore, since *W*_*j*_ = 0 for *j /*∈ *A* and *d < n*_2_,

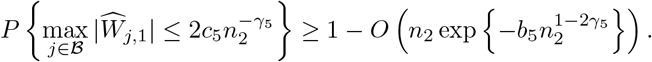

Also, since 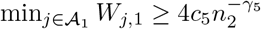,

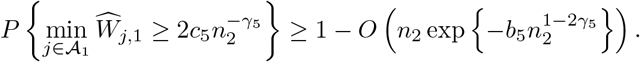

As a result,

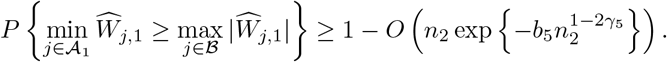

Similarly, for some *b*_6_ *>* 0,

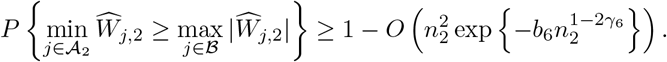

That is, important features are ranked above unimportant ones with probability approaching 1. Given 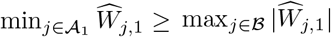 and 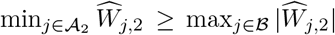, the knockoff procedure stops at 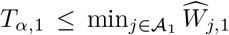 and 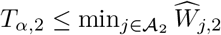 as 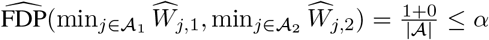, in which case 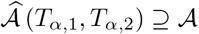.

## 7 Additional Simulation Results

As additional simulation studies, we further consider a linear design with varying signal strength (Example 3) and a more complex nonlinear design (Example 4) for both AFT- and PH-type of models under different censoring mechanisms. **Example 3**. Let **X** ∼ *N*_*p*_(**0**, Σ), where Σ = *AR*(0.5). Given **X**, the true survival time is generated from the following

accelerated failure time (AFT) model and proportional hazard (PH) model:

**1. Model 1:** log *T* = 2*X*_1_ + .8*X*_2_ + .9*X*_3_ + *X*_4_ + 2*X*_5_ + *E*, where *E* ∼ *N* (0, 1) independently;

**2. Model 2:** log(.5(*e*^2*T*^ − 1)) = 2*X*_1_ + .8*X*_2_ + .9*X*_3_ + *X*_4_ + 2*X*_5_ + *E*, where *E* follows the standard extreme value distribution independently.

For each model, the survival time is subject to two censoring mechanisms:

1. independent censoring time *C* generated from uniform distribution on [0, *c*_0_];
2. dependent censoring time *C* generated from exponential distribution with mean *c*_0_*e*^*X*^3,

where the constant *c*_0_ is chosen to achieve 30% or 50% censoring rate (CR). The results are summarized in Table 6 for *n* = 200 and *d* = [*n/* log *n*] = 38.

**Table 6:**
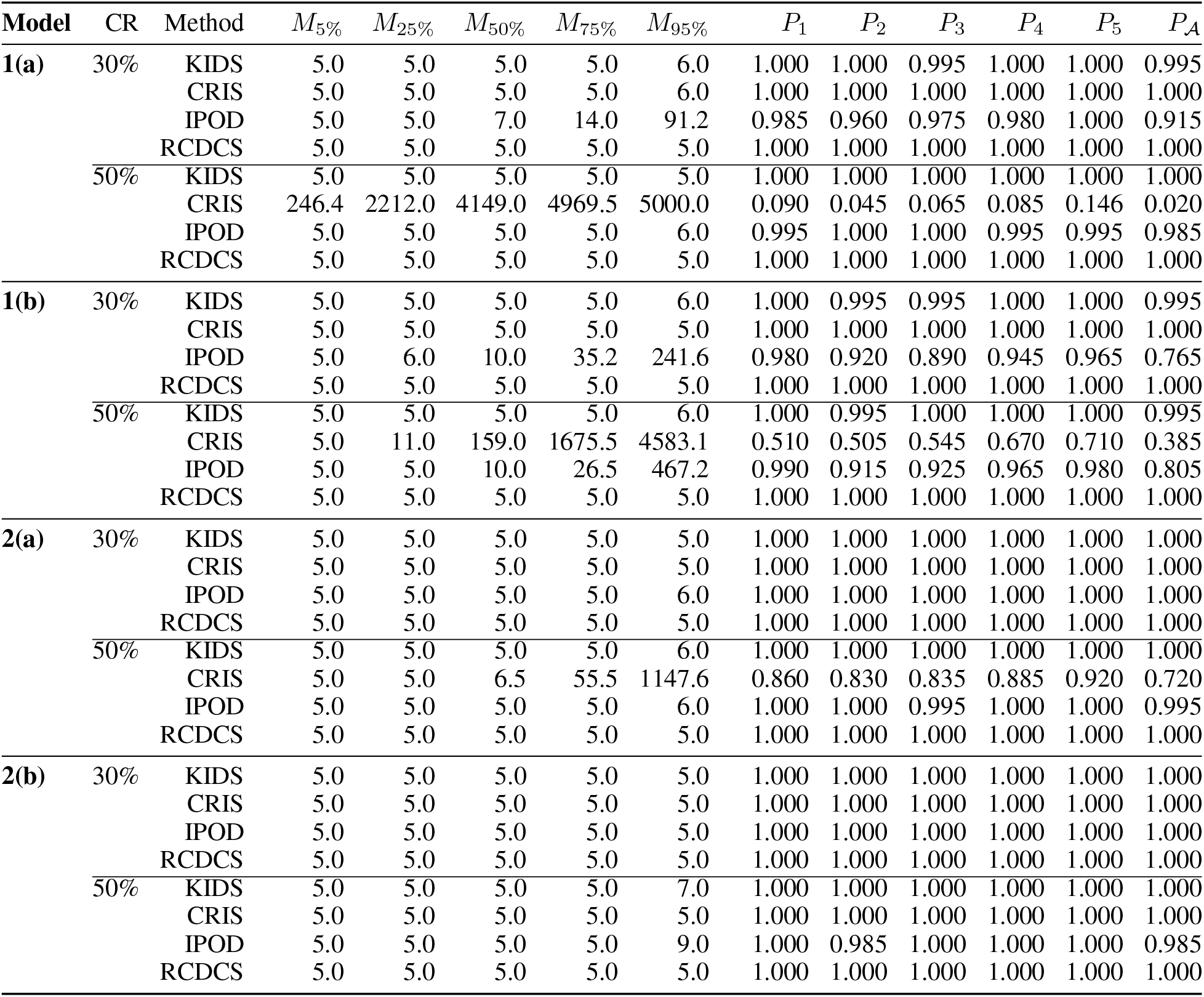
Quantiles of MMS (*M*_*τ*_) and selection proportions (*P*_*j*_’s and *P*_*A*_) for models in **Example 3** based on 200 replicates with *n* = 200, *p* = 5000 and *d* = [*n/* log *n*] = 38.

### Example 4

The setup of this example is identical to Example 3, except that the true survival time is generated from the following accelerated failure time (AFT) model and proportional hazard (PH) model:

**1. Model 3:** log *T* = *g*_1_ + *g*_2_ + *g*_3_ + *g*_10_ + *E*,where *E* ∼ *N* (0, 1) independently and 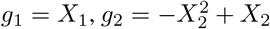,*g*_3_ = 2[*exp*(−3(*X*_3_ − 1)^2^) + *exp*(−4(*X*_3_ − 3)^2^)] and 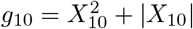:

**2. Model 4:** log(.5(*e*^2*T*^ − 1)) = *g*_1_ + *g*_2_ + *g*_3_ + *g*_10_ + ϵ, where ϵ follows the standard extreme value distribution independently.

The results are summarized in Table 7 for *n* = 200 and *d* = [*n/* log *n*] = 38.

**Table 7:**
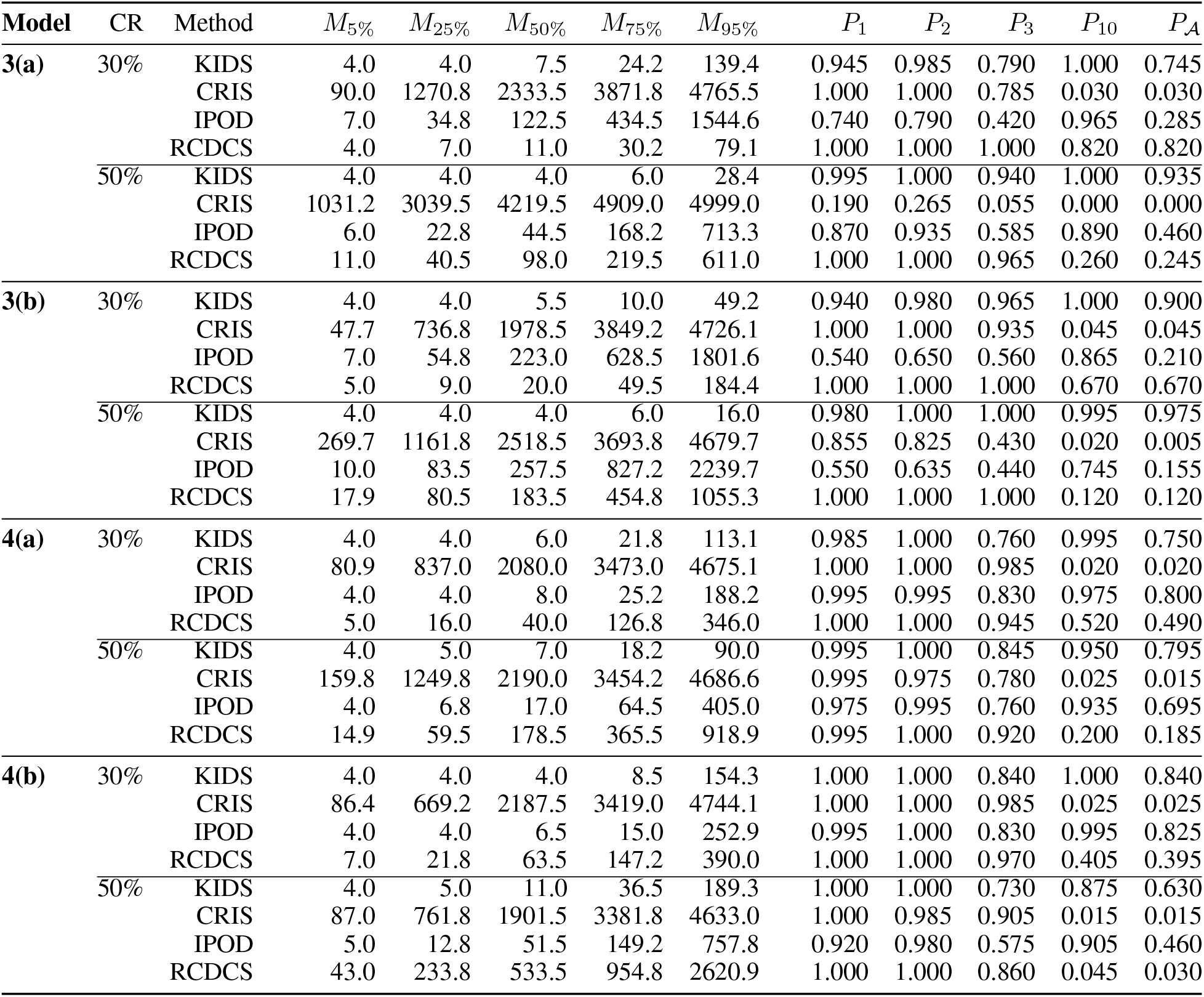
Quantiles of MMS (*M*_*τ*_) and selection proportions (*P*_*j*_’s and *P*_*A*_) for models in **Example 4** based on 200 replicates with *n* = 200, *p* = 5000 and *d* = [*n/* log *n*] = 38.

## 8 Clinical Characteristics of The TCGA and GSE65858 Primary Tumor Samples

A summary of the clinical characteristics of the TCGA and GSE65858 primary tumor samples is presented in Table 8.

**Table 8:** Subgroup frequency (and percentage in parentheses) for clinical characteristics of the TCGA and GSE65858 Primary Tumor Samples.

| Variable | TCGA (n=518) | GSE65858 (n=253) |
| --- | --- | --- |
| Age |  |  |
| $\leq 50$ | 94 (18.15%) | 40 (15.81%) |
| $> 50$ | 424 (81.85%) | 213 (84.19%) |
| Gender |  |  |
| Male | 383 (73.94%) | 210 (83.00%) |
| Female | 135 (26.06%) | 43 (17.00%) |
| Tumor stage |  |  |
| I/II | 100 (22.42%) | 49 (19.36%) |
| III | 81 (18.16%) | 33 (13.04%) |
| IV | 265 (59.42%) | 171 (67.59%) |
| HPV status |  |  |
| Positive | 97 (18.80%) | 73 (28.97%) |
| Negative | 419 (81.20%) | 179 (71.03%) |
| Alcohol history |  |  |
| Yes | 345 (68.05%) | 226 (89.33%) |
| No | 162 (31.95%) | 27 (10.67%) |
| Smoking history |  |  |
| Yes | 389 (76.88%) | 209 (82.61%) |
| No | 117 (23.12%) | 44 (17.39%) |

## Acknowledgements

This research was partially funded by grants P20CA252717, P20CA264067 and R21DE031879 from the United States National Institutes of Health (NIH), and the VCU Quest fund. Services and products in support of the research project were generated by the VCU Massey Comprehensive Cancer Center Biostatistics Shared Resource, supported, in part, with funding from NIH-NCI Cancer Center Support Grant P30CA016059.

